# Environmental impact of Personal Protective Equipment distributed for use by health and social care services in England in the first six months of the COVID-19 pandemic

**DOI:** 10.1101/2020.09.21.20198911

**Authors:** Chantelle Rizan, Malcolm Reed, Mahmood F Bhutta

## Abstract

**Objectives:** Use of Personal Protective Equipment (PPE) has been central to controlling spread of SARS-CoV2. Here we quantify the environmental impact of PPE distributed for use by the health and social care system in England, and model strategies for mitigating the environmental impact.

**Methods:** Life cycle assessment was used to determine environmental impacts of PPE distributed to health and social care in England during the first six months of the COVID-19 pandemic. The base scenario assumed all products were single-use and disposed of via clinical waste. Scenario modelling was used to determine the effect of environmental mitigation strategies; 1) eliminating international travel during supply, 2) eliminating glove use 3) reusing gowns and face shields, 4) maximal recycling.

**Results:** The carbon footprint of PPE distributed during the study period totalled 106,478 tonnes CO_2_e, with greatest contributions from gloves, aprons, face shields, and Type IIR surgical masks. The estimated damage to human health was 239 DALYs (disability adjusted life years), impact on ecosystems was 0.47 species.year (loss of local species per year), and impact on resource depletion was costed at US $ 12.7 (GBP £ 9.3) million.

Scenario modelling indicated UK manufacture would have reduced the carbon footprint by 12%, eliminating gloves by 45%, reusing gowns and gloves by 10%, and maximal recycling by 35%. A combination of strategies may have reduced carbon footprint by 75% compared with the base scenario, and saved an estimated 183 DALYS, 0.34 species.year, and US $ 7.4 (GBP £ 5.4) million due to resource depletion.

**Conclusions:** The environmental impact of PPE is large and could be reduced through domestic manufacture, rationalising glove use, using reusables where possible, and optimising waste management.

## INTRODUCTION

Use of Personal Protective Equipment (PPE) has been a central behavioural and policy response to control spread of the SARS-CoV2 virus during the global COVID-19 pandemic. In particular masks, and sometimes gloves, aprons, gowns, and face/eye protection have been recommended or used in high-risk situations such as healthcare settings or enclosed public spaces. The resultant surge in demand for PPE has required an increase in PPE production, including an estimated 11% increase in global production of gloves in 2020.(1)

Whereas there is evidence that PPE is effective in limiting transmission of the SARS-CoV2 virus, the necessity and extent of PPE for use in different circumstances is still subject to debate.(2) Excessive use of PPE risks generating unnecessary financial cost: for example, UK government budgeted GBP £15bn of funds for purchasing PPE for public sector workers in 2020-21.(3) In addition, use of PPE generates a cost to the environment (which in turn impacts on human health), but to date that risk has not been quantified.

Here we used the approach of life cycle assessment (LCA) to estimate emissions and resulting environmental impact from the most common PPE items prescribed and used in the National Health Service (NHS) and public social care sector in England: masks, gloves, aprons, gowns, and face/eye protection.(4) We equated this with data on the volumes of these products distributed for use by health and social care services in England in the first six months of the COVID-19 pandemic, to estimate the overall environmental impact of PPE over this time period.(5) We evaluated the associated damage to human health (measured in disability adjusted life years), ecosystems (loss of local species), and resource scarcity (financial cost involved in future mineral and fossil resource extraction). We model a number of approaches which could mitigate such impact, and which could inform future policy on use and supply of PPE.

## METHODS

### Selection of representative PPE items and determination of material composition

We based our analysis on products in use at our hospital (Royal Sussex County Hospital, Brighton UK), which is a typical acute tertiary teaching hospital, to represent commonly used PPE. Specifically, we evaluated nitrile gloves, polyethylene aprons, plastic face shields (to represent all eye/face protection), polypropylene fluid-repellent gowns, polypropylene filtering face piece (FFP) respirator masks (both cup fit and duckbill style), Type II polypropylene surgical masks, and Type IIR polypropylene fluid-resistant surgical masks. Type II surgical masks were not available in our hospital setting and so an example was sourced elsewhere, with packaging assumed to be the same as the Type IIR surgical masks.

For each item, we used manufacturer information to determine the raw material composition, or expert knowledge where such information was not available. Each component of the item was weighed using Fisherbrand FPRS4202 Precision balance scales (Fisher Scientific, Loughborough, UK). We included associated primary and secondary packaging up to the packing unit supplied to the hospital.

### Parameters for Life Cycle Assessment

A LCA was conducted in accordance with ISO 14044 Guidelines,(6) and modelled using SimaPro Version 9.10 (PRé Sustainability, Amersfort, Netherlands), with additional analysis using Microsoft Excel for Mac Version 16.25 (Microsoft Corp, Washington, US). We performed a ‘cradle to grave’ LCA, including raw material extraction, manufacture, transport, and disposal (system boundary outlined in Supplementary Figure 1).

The previously determined composition and weight of materials in each item and packaging was matched with closest materials within the Ecoinvent database Version 3.6 (within SimaPro), to determine material specific global average impacts of raw material extraction, production, and transport to the ‘end user’ (in this case the manufacturer).

For manufacturing, the country of origin was modelled based on those reported by the NHS PPE Dedicated Supply Channel.(7) Where a type of product was procured from more than one country, a weighted average was applied to our calculations on the assumption that equal numbers of the product were distributed from each listed supplier.(7) A list of suppliers awarded UK contracts for PPE is available in a UK National Audit Office report, but this does not provide data on volumes supplied or distributed, or country of origin,(3) and our model was based on best available information on the country of origin of products.(7) Electricity consumption during the manufacturing process was modelled on best available secondary data for comparable products,(8-11) and electricity inventory processes chosen and weighted based on country of origin(s). We excluded water and fuel during manufacture because such data were not available, and are unlikely to materially affect results.

For transportation we assumed all items were shipped from the country of origin to the UK, because this method of transport was most commonly used (>80%) for PPE from new suppliers to the UK during the period included in the study.(3) We included 160km of travel by road via heavy goods vehicles both within the country of origin and in the UK, with an additional 8km at either end of each journey via courier. All transportation distances were estimated using the online Pier2Pier tool (Supplementary table 1).(12)

The processes in relation to raw material extraction, manufacture, transport, and disposal selected for the LCA inventory within SimaPro are shown in Supplementary table 2. We modelled life cycles on the basis that all items were used only once, and (in accordance with UK guidance) disposed of as clinical waste(13) via high-temperature hazardous incineration.(14)

### Impact assessment methodologies

Following development of the LCA inventory within SimaPro, we used the World ReCiPe Midpoint Hierarchist method Version 1.1 (integrated within SimaPro) to characterise emissions from the lifecycle inventory assessment and to combine these into environmental impacts. ReCiPe is a widely used impact assessment method, chosen because it considers a broad range of global environmental impacts (unlike others which are regional), and because it considers impacts at two levels; midpoint and endpoint. Eighteen midpoint impact categories (considering single environmental problems) are evaluated using this method: global warming, stratospheric ozone depletion, ionising radiation, ozone formation (on human health, and terrestrial ecosystems), fine particulate matter formation, terrestrial acidification, eutrophication (freshwater, and marine), ecotoxicity (terrestrial, freshwater, marine), toxicity to humans (relating to chemicals with reported carcinogenic [cancer] effects such as polyaromatic hydrocarbons, and chemicals with non-carcinogenic effects [non-cancer diseases] such as lead, which is associated with learning disabilities at high blood levels), land use, resource scarcity (mineral, and fossil), and water consumption. Global warming was the primary impact evaluated, with greenhouse gases summated and expressed as carbon dioxide equivalents (CO_2_e), providing a ‘carbon footprint’. We also aggregated midpoint impact categories to calculate endpoint factors for damage to human health, the natural environment, and resource scarcity, as per the ReCiPe Endpoint Hierarchist method, Version 1.1.

We calculated environmental impact values per item, and then multiplied by the total number of PPE items supplied to health and social care services in England between 25^th^ February and 23^rd^ August 2020, using publicly available volumes data.(5) For respirator masks we combined volumes of FFP2 and FFP3 masks, and assumed an equal split between cup fit and duckbill styles. Table 1 details extracted parameters on electricity in manufacture, and on country of origin, and volumes of PPE distributed for use by health and social care services in England. We used ReCiPe Hierarchist method normalisation factors to compare our calculated total midpoint and endpoint impacts to the mean average contributions to each of those impacts from a global average person’s daily routine activities over a six month period.

**Table 1:** Model parameters for manufacture and supply of PPE items. Model parameters and source of data on electricity within manufacture (not including production of material), country of origin, and number distributed to health and social care services in England (25^th^ February-23^rd^ August 2020). FFP=filtering facepiece.

| **Product** | **Manufacturing electricity**<br>**(kWh/kg product)** | **Country of origin<sup>7</sup>**<br>**(ratio)** | **Number distributed**<br>**Feb-Aug 2020<sup>5</sup>** |
| --- | --- | --- | --- |
| Apron | 0.490<sup>10</sup> | China, Thailand (9:1) | 441,061,000 |
| Face shield | 0.367= mean<sup>8,10,11</sup> | China (1) | 45,326,000 |
| FFP respirator (any type) | 0.296<sup>11</sup> | China, UK, France (1:1:1) | 37,212,000 |
| Gloves (single) | 2.790<sup>9</sup> | Malaysia (1) | 1,839,235,000 |
| Single-use gown | 0.316<sup>8</sup> | China, Egypt, Germany (9:4:3) | 5,985,000 |
| Surgical mask (Type II) | 0.296<sup>11</sup> | China, UK, Mexico (11:2:1) | 6,623,000 |
| Surgical mask (Type IIR) |  |  | 479,341,000 |
| Disinfectant wipe | 0.296<sup>11</sup> | China (1) | - |
| Reusable gown | 0.316<sup>8</sup> | China, UK (3:1) | - |

### Scenario modelling

We modelled the effect of four approaches that could mitigate the environmental impact of PPE manufacture, supply and disposal.

First, we modelled the impact of domestic (UK) manufacture of products, effectively eliminating international transport (shipping) but using the same road travel assumptions, with UK electricity grid inventory process data for manufacture.

Second, we modelled reducing glove use by replacing use of gloves (with subsequent hand washing), with hand washing alone (which can effectively destroy the virus).(15) It was not necessary to calculate the environmental impact of hand washing itself, because hand washing is common to both scenarios, and so not relevant to comparative analysis.

Third, we modelled the impact of using reusable gowns and reusing face shields. Reusable gowns were assumed to be laundered and re-used 75 times before disposal (based upon direct correspondence with the manufacturer). We extracted energy, water, and detergent requirements, based upon previously published studies and reports.(8, 16) Detergent chemical composition was included where known, and where the chemical constituted ≥1% of the detergent composition (Supplementary table 3). The transportation of linen from the user site to a laundering facility was assumed to be 160 km (round trip) via heavy goods vehicles by road. Face shields were assumed to be reused five times, with cleaning by a disinfectant wipe between uses-an accepted practiced in the UK.(17) To inform this model we calculated the environmental impacts of reusable gowns and disinfectant wipes, using the same approach detailed earlier for other PPE (except that secondary packaging of reusable gowns was excluded, as this was likely to reach the insignificance threshold of contributing <1% to the impact).(18) The country of origin of the reusable gown and disinfectant wipes in use at our hospital were assumed to be representative, and determined using either the packaging or direct contact with the manufacturer.

Fourth, we modelled the environmental impact of maximal recycling of products, assuming it was possible to recycle all items and their components. We used the open-loop ‘recycled content method’, which allocates subsequent emissions and environmental impacts of the recycling process, and net reduction of virgin material acquisition, to the production of the recycled goods.(18)

Finally, we modelled the environmental impact of combining these mitigation measures.

We also modelled the effect of changes that could increase the environmental impact, specifically a change of overseas transportation to air freight. Air freight was employed for rapid delivery of PPE supplies to the UK early in the COVID-19 pandemic because of insufficient stock.(19)

## RESULTS

The composition and weight of materials for each item of PPE and associated packaging are detailed in Table 2.

**Table 2:** Material composition of PPE. All items single-use aside from components of reusable gown. FFP=filtering facepiece, LDPE= low density polyethylene, no.= number, *= assumed proportion

| Product | Example manufacturer and model | Item Material | Weight (g)<br>(no. uses when >1) | Packaging Material | Weight (g) | Number of items/<br>package |
| --- | --- | --- | --- | --- | --- | --- |
| Apron | Trans-Continental Marketing<br>TCMAPR200W | LDPE film | 9.86 | LDPE film<br>Corrugated cardboard | 6.36<br>365.67 | 200<br>1000 |
| Face shield | Miers<br>Mio-shield | LDPE film<br>Polyurethane foam<br>Synthetic rubber | 18.53<br>5.44<br>4.95 | LDPE film<br>Corrugated cardboard | 8.48<br>683.57 | 1<br>200 |
| FFP respirator<br>(cup fit) | 3M<br>FFP3 8833 | Non-woven polypropylene<br>Melt-blown polypropylene<br>Polypropylene injection moulded<br>Polyurethane foam<br>Synthetic rubber<br>Aluminium<br>Steel | 5.98<br>2.99<br>3.93<br>2.54<br>2.55<br>1.13<br>0.32 | Boxboard<br>Corrugated cardboard | 88.32<br>350.40 | 20<br>120 |
| FFP respirator<br>(duckbill) | 3M<br>FFP3 1863 | Non-woven polypropylene<br>Melt-blown polypropylene<br>Polyurethane foam<br>Synthetic rubber<br>Steel | 5.52<br>2.76<br>0.85<br>0.77<br>0.48 | LDPE film<br>Boxboard<br>Corrugated cardboard | 1.25<br>88.32<br>350.40 | 1<br>20<br>120 |
| Gloves<br>(single) | Schottlander<br>Soft touch flexible nitrile | Synthetic rubber | 3.23 | Boxboard<br>Corrugated cardboard | 64<br>382.55 | 200<br>1000 |
| Single-use gown | YADU<br>Medical surgical gown reinforced | Non-woven polypropylene<br>Synthetic rubber | 134.74<br>7.72 | LDPE film<br>Paper<br>Corrugated cardboard | 4.43<br>6.46<br>911.22 | 1<br>1<br>50 |
| Surgical mask (Type II) | Genmed Surgical face mask Type II elastic strap | Non-woven polypropylene<br>Melt-blown polypropylene<br>Synthetic rubber<br>Aluminium | 1.00<br>0.50<br>0.28<br>0.23 | Boxboard<br>Corrugated cardboard | 41.75<br>843.30 | 2000<br>50 |
| Surgical mask (Type IIR) | Arma Pennine Disposable medical face mask 3-ply with earloops | Non-woven polypropylene<br>Melt-blown polypropylene<br>Synthetic rubber<br>Aluminium | 1.63<br>0.82<br>0.45<br>0.20 | Boxboard<br>Corrugated cardboard | 41.75<br>843.30 | 50<br>2000 |
| Disinfectant wipe | Clinell Universal Wipe | Non-woven polyester<br>Quaternary ammonium compounds and polymeric biguanide hydrochloride | 2.28<br>0.02* | Polypropylene, oriented film<br>Polypropylene, injection moulding<br>Corrugated cardboard | 13.28<br>10.90<br>406.47 | 200<br>200<br>1200 |
| Reusable gown | Elis Standard protection reusable gown | Polyester microfibre<br>Synthetic rubber<br>Tissue paper | 301.37(75)<br>18.31(75)<br>13.41 | LDPE film<br>Paper | 6.42<br>19.90 | 1<br>1 |

### Impact assessment: the environmental impact of PPE

The carbon footprints of individual items were estimated as follows: single-use gowns 905 g CO_2_e, face shield 231 g CO_2_e, cup fit FFP respirator 125 g CO_2_e, duckbill FFP respirator 76 g CO_2_e, apron 65 g CO_2_e, single glove 26 g CO_2_e, Type IIR surgical mask 20 g CO_2_e, and Type II surgical mask 13 g CO_2_e. The mean contribution of production of materials to the overall carbon footprint of items was 46% (range: 35 - 49%), 39% from clinical waste (range: 32 - 40%), 6% from production of packaging materials (range: 0.5 – 16%), and 5% from electricity used within manufacturing (range 2 - 29%), and 4% from transportation (range 3 - 6%) (Figure 1). Supplementary tables 4-11 show contributions per item across all environmental impacts, with further breakdown of processes. Supplementary figures 2-9 provide network diagrams visualising the process drivers of global warming impact.

**Figure 1:**
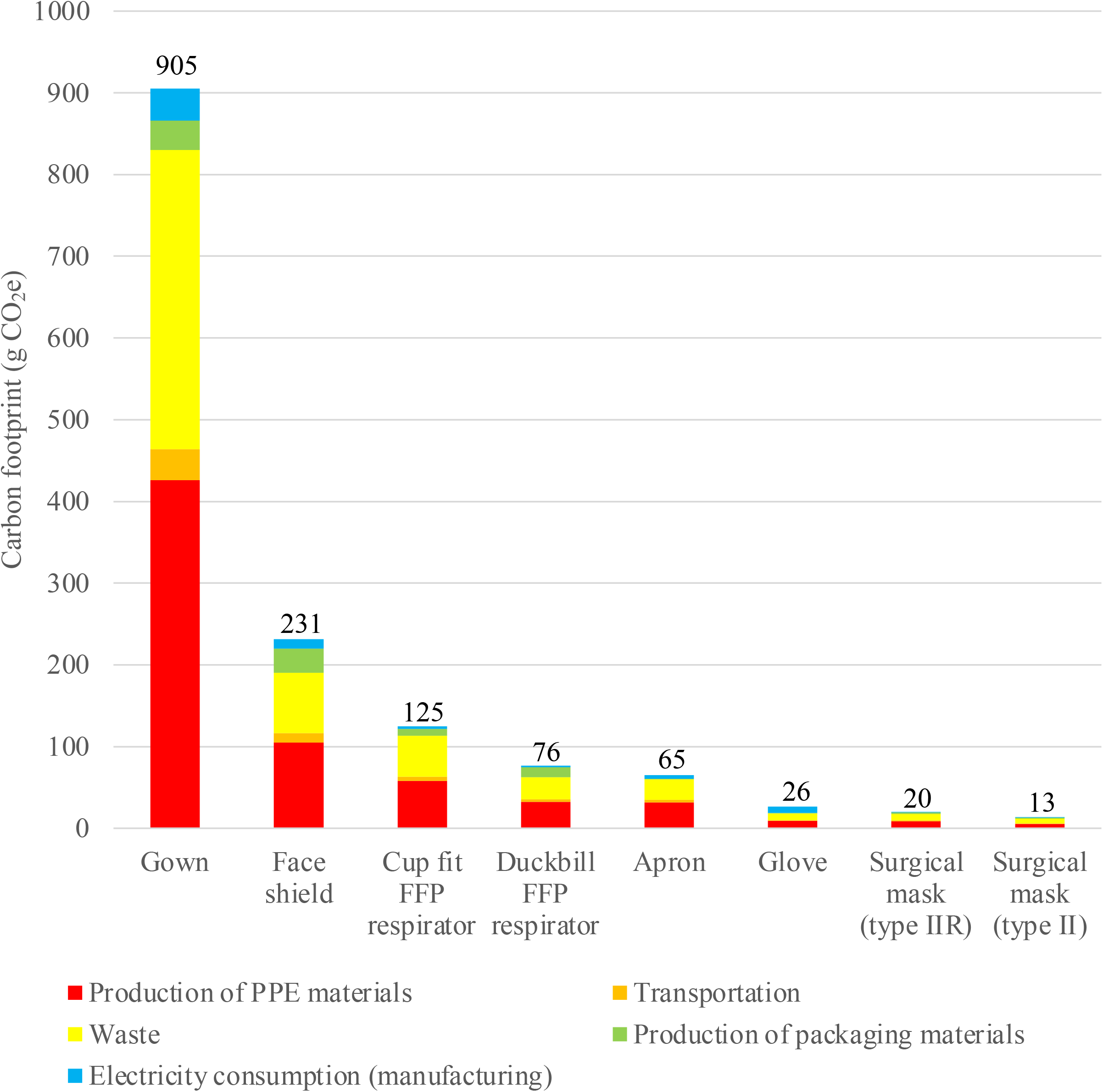
Carbon footprint of individual PPE items. Carbon footprint of individual single-use PPE items, with breakdown of process contributions. Production of X materials includes the raw material extraction, production, and transport to the PPE manufacturer. CO_2_e= carbon dioxide equivalents, FFP= filtering facepiece

The carbon footprint of all PPE supplied to health and social care services in England between 25^th^ February-23^rd^ August totalled 106,478 tonnes CO_2_e. This equated to 26,662 times the global average person’s carbon footprint during a six-month time period (normalised results Table 3). The proportional contribution for each type of item was primarily determined by the volumes distributed of that item, and was greatest for gloves, followed by aprons, face shields, and Type IIR surgical masks. The relative impact of PPE distributed during this period on the carbon footprint and other midpoint environmental measures are detailed in Table 3. Endpoint impact results estimated that the total damage to human health during this period was 239 DALYs (disability adjusted life years), equating to 20,126 times the average person’s contribution to DALYs over a similar period. The impact on ecosystems was 0.47 species.year (loss of local species per year), equivalent to 1,300 times a person’s impact in this category over the study period. The impact on resource depletion equated to US $ 12.7 (GBP £ 9.3) million involved in future mineral and fossil resource extraction, equal to 907 times a person’s average contributions to resource depletion over six months.

**Table 3:**
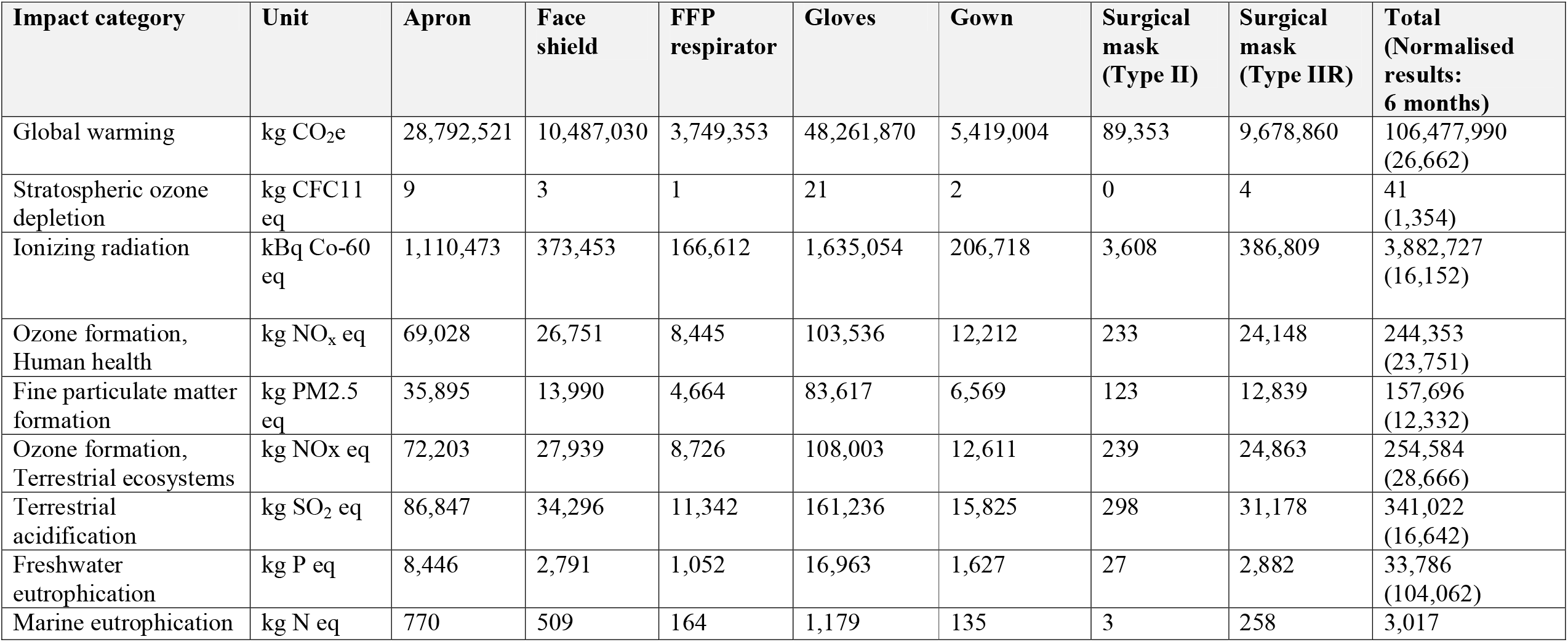
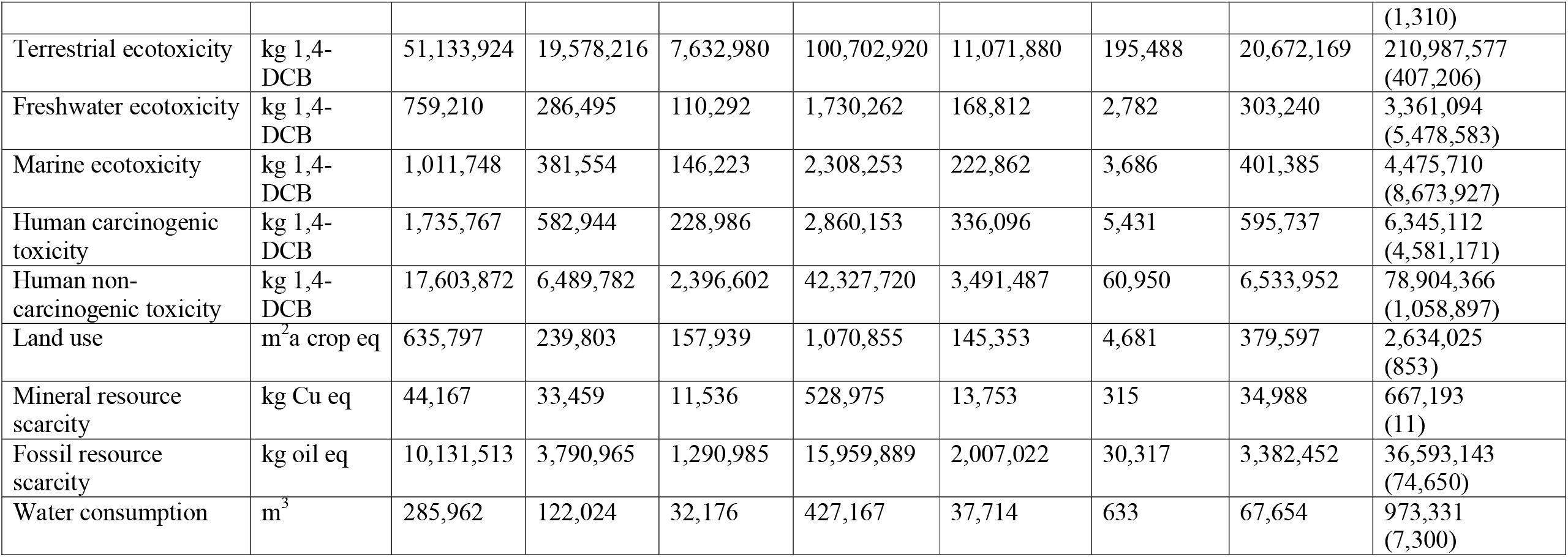
Environmental impact of PPE distributed to health and social care services in England Feb-Aug 2020. Environmental impacts (midpoint categories) measured using life cycle assessment and modelled on total volumes of core PPE distributed to health and social care services in England between 25^th^ February and 23^rd^ August 2020. Normalised results= environmental impact relative to the global average person’s contribution to the impact category over 6 months. For example, a normalised figure of 26,662 for the global warming category indicates that the greenhouse gas emissions relating to the total volume of PPE supplied during the study period equates to the total amount of greenhouse gases expected to be generated by the activities of 26,662 average people over the course of six months. FFP = filtering facepiece,1,4-DCB =dichlorobenzene, CFC11= Trichlorofluoromethane, CO_2_e= carbon dioxide equivalents, Cu= copper, eq= equivalents, FFP= filtering facepiece, kBq Co-60 eq = kilobecquerel Cobalt-60, FFP= filtering facepiece, m^2^a = square meter years, N= nitrogen, NO_x_= nitrous oxides, P=phosphate, PM2.5 = particulate matter <2.5 micrometers, SO_2_= sulphur dioxide

### Scenario analysis: mitigating the environmental impact of PPE

The carbon footprint of PPE was reduced by 12% through manufacturing PPE in the UK, saving 12,491 tonnes CO_2_e over the six month study period. Reductions were due to the elimination of overseas travel (2.4%), alongside use of UK electricity (9.3%), (which has a higher proportion of renewables compared with the majority of countries of origin assumed in the base scenario).

Eliminating glove use would have reduced the carbon footprint by 45%, saving 48,262 tonnes CO_2_e over 6 months.

For reuse, the environmental impact of one use of a reusable gown was lower than that of a single-use gown across 16/18 environmental midpoint impact categories (with impact reductions of 17% to 86%) (Supplementary figure 10). The impact of reusable gowns on marine eutrophication was 47% greater than single-use gowns, with 72% of this impact from wastewater generated during the laundering process (Supplementary figure 11). The impact of reusable gowns on land use was more than double that of single-use gowns, with 86% of this effect due to single-use paper within the associated hand-towel and packaging (Supplementary figure 12). Reusing face shields five times with disinfectant wipe between use showed 57% to 73% lower impact across all midpoint categories, when compared with single use (Supplementary figure 13). Opting for reusable gowns and reusing face masks could have saved 11,107 tonnes of CO_2_e over the study period (10% of the total for all PPE distributed).

Maximal recycling reduced the carbon footprint of PPE by 35% (saving 37,266 tonnes CO_2_e).

A combination of UK manufacturing, eliminating glove use, reuse of gowns and face shields, and maximal recycling could have led to a 75% reduction (saving 79,830 tonnes CO_2_e) (Figure 2). Results of other midpoint impact categories are detailed in Supplementary table 12.

**Figure 2:**
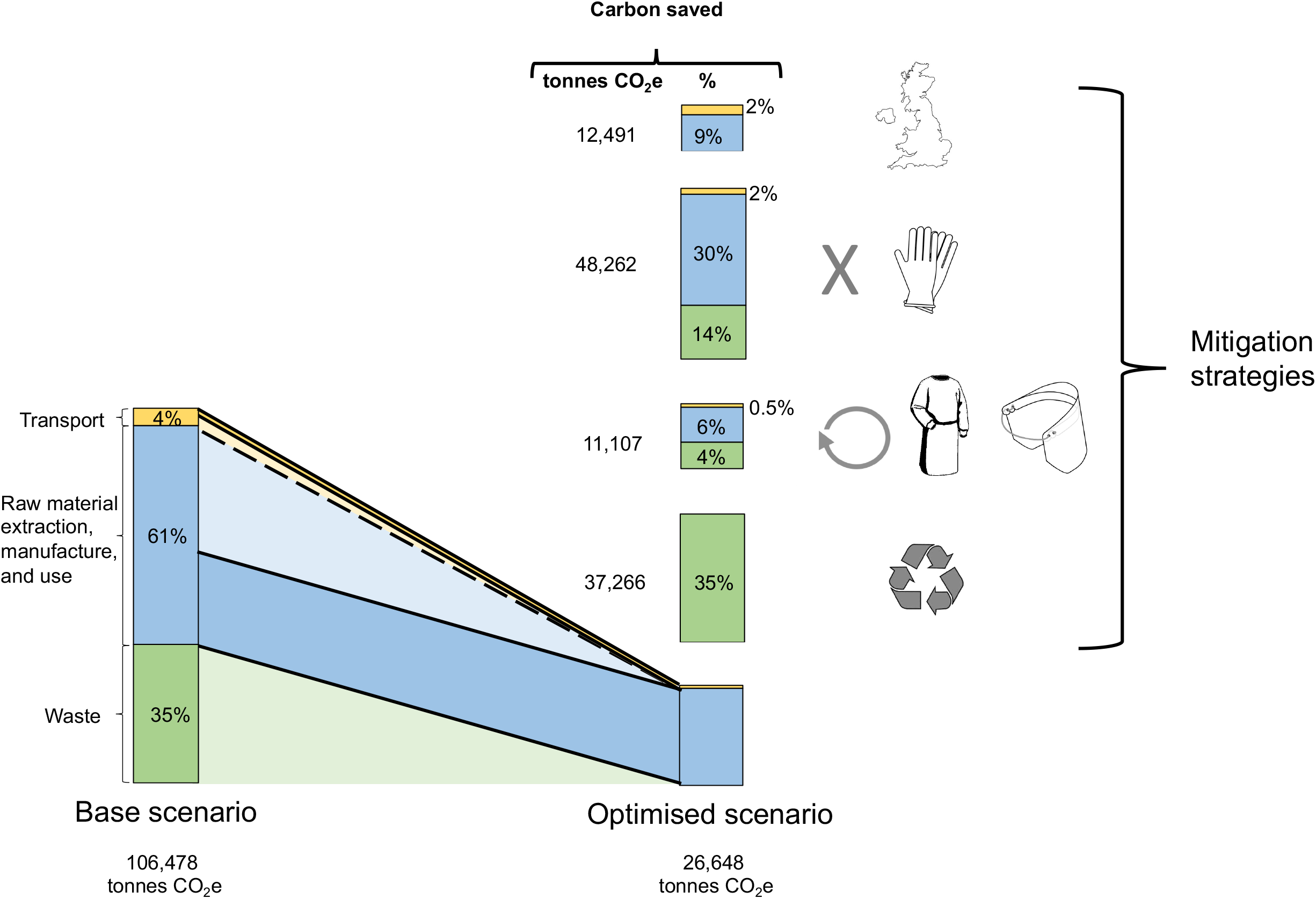
Mitigating the carbon footprint of PPE. Bar graph to left demonstrates the carbon footprint of the base scenario PPE use (modelled on total volumes of core PPE distributed to health and social care services in England between 25^th^ February and 23^rd^ August 2020, and assuming shipping, single-use PPE and clinical waste), totaling 106,478 tonnes CO_2_e (carbon dioxide equivalents). Bar graph to right at base demonstrates mitigation of the carbon footprint (modelled through combining scenarios), totaling 26,648 tonnes CO_2_e. Bar graphs stacked above optimised scenario demonstrate the carbon savings from each mitigation strategy (reported in tonnes and as a percentage, compared with the base scenario). Scenarios from top to bottom: UK manufacture, eliminating glove use, reuse of gowns and face shields, recycling.

Endpoint category scenario modelling showed a similar pattern, with largest reductions seen through eliminating glove use (Figure 3, Supplementary table 13). Maximum reductions through a combination of UK manufacture, eliminating glove use, reusable gowns and face shields, and maximal recycling would have saved an estimated 183 DALYS, 0.34 species.year, and US $ 7.4 million (GBP £ 5.4 million) due to resource depletion.

**Figure 3:**
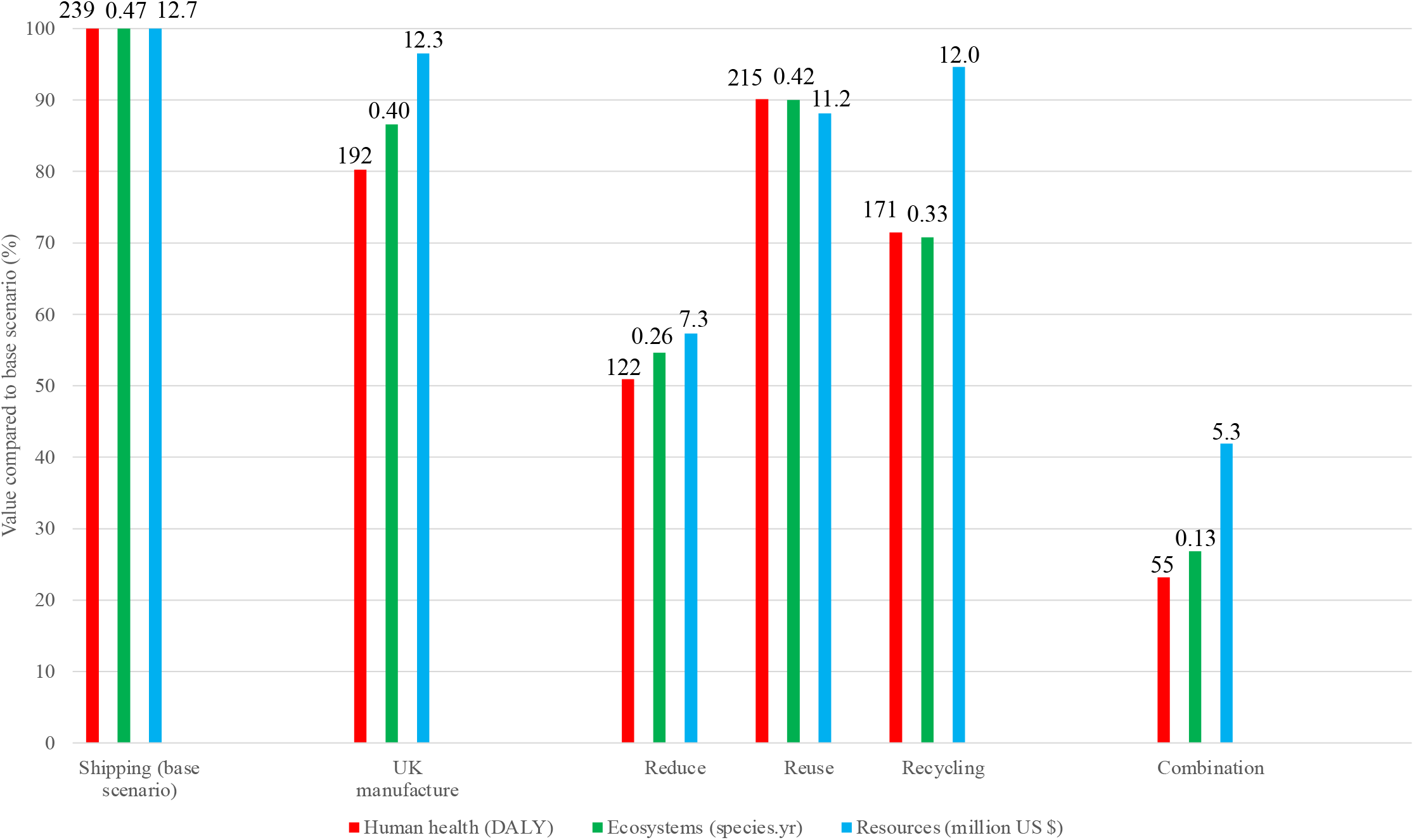
Environmental impacts of alternative scenarios. Environmental impacts (endpoint categories) of alternative scenarios, modelled on total volumes of core PPE distributed to health and social care services in England between 25^th^ February and 23^rd^ August 2020, normalised to highest scenario for each impact factor, modelling base scenario (shipping, single-use PPE, clinical waste), use of UK manufacture, shipping, reduce (zero glove use), reuse (reusable gown, reuse of face shield, all other items single-use), recycling, and combination of measures. Data labels above bars relate to absolute values, measured in disability adjusted life years (DALYs), loss of local species per year (species.year), and extra costs involved for future mineral and fossil resource extraction (US $).

### The effect of air freight on environmental impact

Use of air freight in place of shipping increased the carbon footprint of PPE by 50%. This would have increased the carbon footprint by 52,360 tonnes CO_2_e over the six month study period, and resulted in an additional 75 DALYS, 0.20 species.year, and US $ 7.7 (GBP £ 5.6) million due to resource depletion (compared with base scenario).

## DISCUSSION

We estimate the carbon footprint of PPE distributed for use by health and social care services in England during the first six months of the COVID-19 pandemic to be 106,478 tonnes CO_2_e, which is equivalent to 0.8 % of the entire carbon footprint of health and social care in England during six months of normal activity (estimated at 27 million tonnes CO_2_e per annum in 2018).(20) Per day this equates to a mean of 591 tonnes CO_2_e, equivalent to 27,000 times the average individual’s carbon footprint, or around 244 return flights from London to New York.(21)

There are some caveats in interpretation of these data. Around 3 billion items of PPE were used in the six-month period analysed, but data from 2019 (prior to the pandemic) suggest that around 1.2 billion items would normally be consumed in the NHS in a six-month period,(5) hence the excess in this period was in fact 1.8 billion items. However, 70-80% of elective care in the NHS stopped during the first few months of the pandemic, and emergency attendances decreased by 30-40%,(22) meaning PPE use related to normal (non-pandemic) NHS activity would have fallen by a comparable amount. The backlog of pending elective activity means that the PPE required to deliver elective care has been deferred rather than abolished, and as elective activity resumes, current requirements for enhanced protection mean that use of PPE will likely be even higher than before.

The large environmental impact of PPE, and the probability that we will continue to require and use high volumes of PPE for the foreseeable future, demands an urgent evaluation of approaches to reduce this impact.

### Opportunities to mitigate the environmental harm of PPE

Strategies to mitigate environmental impact are often based on principles of reduce, reuse and recycle, and we believe this approach can also be applied to PPE, and without compromising safety.

In healthcare settings in England the policy at the time of the study period mandated use of gloves for close patient contact,(4) although transmission of coronavirus is thought to occur mostly via airborne spread rather than direct transfer.(23) Hand washing can destroy the SARS-CoV2 virus,(15) so may negate the need to wear gloves. In our six-month analysis period, nearly 1.8 billion gloves were distributed to health and social care services, and these volumes increased in July and August 2020(5) despite a reduction in the number of COVID-19 cases in the same time period. Gloves accounted for 45% of the total carbon footprint of PPE in our study, so a policy to rationalise glove use could have a large impact on environmental harm. Furthermore, aprons accounted for 27% of the carbon footprint of PPE, and there may be parallel opportunities for policy change to reduce use of aprons, again without compromising safety. Current UK guidance advises that gloves and aprons are no longer required where contact with patients is minimal, although the existence of contradictory local policy documents(24) suggests that these may not have been universally adopted. The UK government also supports reducing glove use because of their association with contact dermatitis amongst healthcare staff.(25)

Where PPE is required, our data suggest use of domestic manufacture could reduce associated carbon footprint by 12%. Domestic PPE manufacture has been used in many countries in response to PPE shortages, including Germany, where the government introduced a scheme to support German manufacture of facemasks.(26) The UK government has released a policy seeking to develop and maintain a domestic manufacturing base to improve resilience,(3) with the ambition that this will meet the majority (70%) of PPE demands, although this target excludes gloves which are responsible for a large proportion of PPE.(25) The policy also includes adequate stockpiling of PPE, and this, together with domestic manufacture, will mitigate the need to urgently air freight PPE from abroad with the associated 50% increase in carbon footprint.

Reuse of PPE is also feasible, and practised in some settings. Extended use of masks is supported by several guidelines(27) and should be encouraged to reduce environmental impact. Reprocessing by sterilisation through chemical or physical means has also been explored,(28) although not widely implemented. For users who wear a mask often, reusable passive or powered air purifiers may have a lower overall carbon footprint, although that was not formally evaluated here. Face shields are already reused in many settings, and our analysis suggests that cleaning with disinfectant wipes and reusing five times lowers carbon footprint by 70% compared to single-use. Reusable gowns are already available and utilised in operating theatres, and we found use of reusable rather than disposable gowns would reduce carbon footprint by two-thirds (consistent with calculations by other authors).(8) To provide reusable gowns to ward or outpatient settings would need an increase in stock, upscaling of laundering facilities, and ideally removal of the hand towels and inner wrap typically supplied with gowns used in operating theatres. The UK government has released policy supporting manufacture of reusable PPE alternatives where possible (including eye protection and gowns), and reprocessing of single-use PPE in emergency circumstances (for example moist heat treatment or hydrogen peroxide vapour for FFP3 masks).(25)

Maximal recycling reduced carbon footprint by 35%, but is unrealistic because adequate infrastructure for waste segregation does not currently exist, particularly for multi-component products such as masks (which would require disassembly of potentially infected materials). However, if PPE in the NHS was disposed of via infectious waste streams rather than clinical waste streams (where it may be decontaminated prior to disposal through recycling, landfill, or low temperature incineration with energy from waste),(14) then the carbon footprint of disposal could be at least halved.(29) A recent LCA study modelled alternative disposal of PPE, finding that decentralised (local) incineration was preferrable to centralised incineration or landfill across all environmental impacts assessed, and opportunities to reduce transportation of waste to such facilities should be explored.(30)

We recognise that a complete implementation of these strategies is not possible or practical, but the models we provide do suggest where policy changes could have impact. The extent to which each of the environmental mitigation strategies can be implemented in practice will be dynamically dependant on multiple factors including industry response to the call for greater domestic manufacture of PPE, availability of local facilities for recycling, and local policies and attitudes. This warrants closer examination, but is beyond the scope of this paper.

### Limitations to our dataset

Our calculations are based on a number of assumptions. We calculated the environmental impact on a single typical model of each type of PPE, but there will be other models and brands, with differences in environmental harm. However, the differences are likely small and unlikely to make substantive changes to the estimates of total environmental impact of the PPE distributed, relative contributions from different product types, or magnitude of mitigation strategies. We assumed all waste was disposed of via clinical waste as recommended, but in reality some PPE incorrectly enters other hospital waste streams (which have a lower environmental impact). We assumed air freight for all PPE from outside the UK, but we only included a single direction of air transport and some PPE may have been shipped.

The actual quantity (and so carbon footprint) of PPE used in health organisations in England during the pandemic is larger than we have included here. Our estimates do not include PPE procured outside of the government dedicated supply channel, including gloves or gowns for use in the operating theatre (which are procured through different channels), or PPE procured by private organisations. We found no publicly available data on PPE procured in other countries, although one supplier in the USA (Project N95) records over 2 million items of PPE supplied over six-months from mid-May to November 2020.(31)

### Looking beyond our dataset

Outside of the healthcare setting, other organisations and individuals will procure PPE, particularly following new policy in countries such as the UK for the use of masks when indoors.(32) In our data the majority of PPE was manufactured partially or completely from plastics or petroleum-based synthetic rubbers, including for example nitrile for gloves, polypropylene for masks and gowns, and polyethylene for visors and aprons, which we estimated to have a mass of over 14,000 tonnes over the six-month period. Disposal of PPE outside of healthcare will mostly be in domestic waste streams which may enter landfill, risking plastic pollution. Discarded masks and gloves have been found polluting oceans.(33)

We are also aware of social (alongside environmental) harms from PPE. There have been longstanding concerns about abuse of workers manufacturing masks and gloves,(34) and such concerns have continued or been exacerbated in recent months, with reports of forced labour to make masks in China,(35) and abuse of migrant workers in factories producing gloves in Malaysia.(36)

## Conclusion

The environmental impact of PPE is substantial and requires urgent review to mitigate effects on planetary health. The most opportune and impactful strategies may be through reduced use of gloves by using hand washing alone, domestic manufacture of PPE, and extended use or reuse of PPE such as masks and gowns. These possibilities warrant further investigation and analysis of feasibility and safety, as well as engagement of policy makers around the globe.

## Data Availability

Supporting data is available in supplementary files

## Ethical approval

Not required- this study did not involve any patients or healthcare staff.

## Guarantor

Chantelle Rizan is the manuscript guarantor.

## Contribution statement

All authors fulfilled the authorship criteria as follows:

- Substantial contributions to the conception or design of the work; or the acquisition, analysis, or interpretation of data for the work; AND
- Drafting the work or revising it critically for important intellectual content; AND
- Final approval of the version to be published; AND
- Agreement to be accountable for all aspects of the work in ensuring that questions related to the accuracy or integrity of any part of the work are appropriately investigated and resolved.

CR led on LCA study design and analysis, and co-authored the first draft of the manuscript. MB initiated the article, sourced sample PPE materials, and co-authored the first draft of the manuscript. MR provided critical input to structure and content. The corresponding author attests that all listed authors meet authorship criteria and that no others meeting the criteria have been omitted.

## Acknowledgements

None to declare

