## Supplementary Figure 1 for "Environmental impact of Personal Protective Equipment distributed for use by health and social care services in England in the first six months of the COVID-19 pandemic"

**Supplementary Figure 1: system boundary**

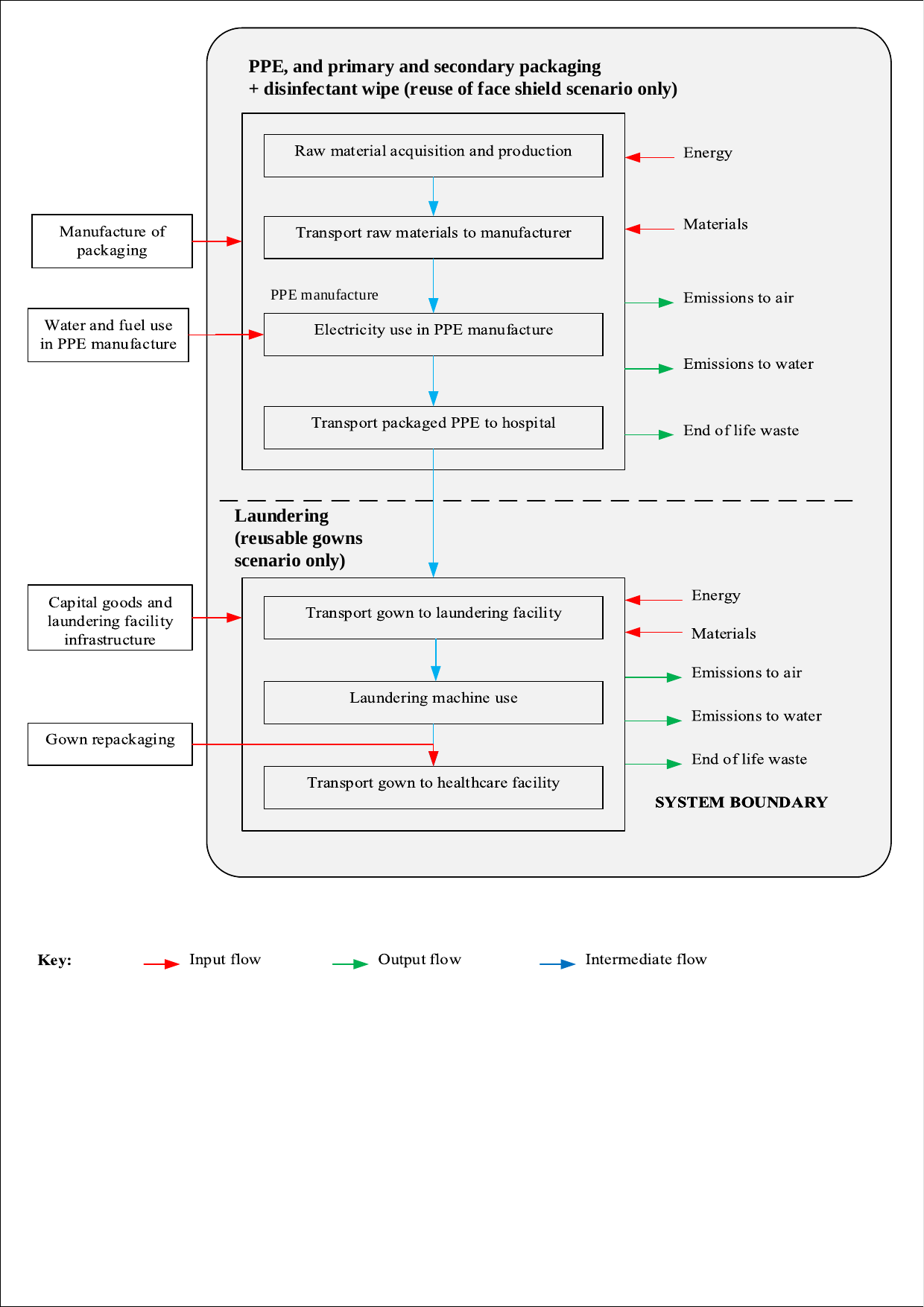

System boundary for life cycle assessment (LCA) of Personal Protective Equipment (PPE). Processes included (for PPE and associated packaging) were energy and materials required for raw material acquisition, production and transport, alongside electricity used in PPE manufacture. Water and fuel use during manufacture of PPE, and the manufacture of associated packaging were excluded. The same processes were included for the detergent wipe modelled in the reuse of face shields scenario. Processes included in laundering (for the reusable sterile gowns scenario only), were energy and materials required by the laundering machine, and associated transport. Capital goods and laundering facility infrastructure were excluded, alongside gown repackaging. End of life waste was included for all materials.

**Supplementary Figure 2: Network diagram for apron showing global warming impact drivers (5% cut-off)**

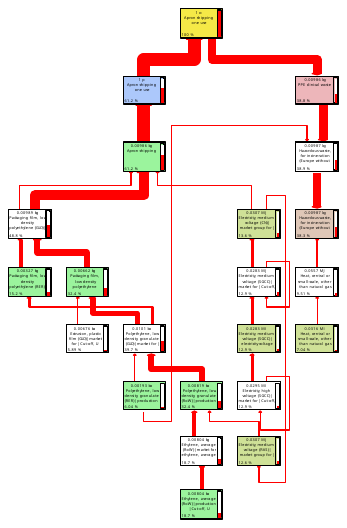

Network diagram for LCA of a single apron, showing drivers for the global warming impact category. Each box represents a unit process (only those >5% contribution to the impact are shown), with percentage contribution shown in bottom left, and quantity of the process for the assembly at the top of each box. The arrows represent the flow of materials between processes, and their thickness reflect the relative contribution.

**Supplementary Figure 3: Network diagram for face shield showing global warming impact drivers (5% cut-off)**

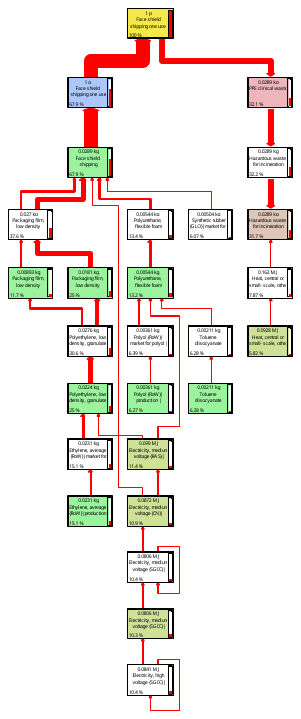

Network diagram for LCA of a single face shield, showing drivers for the global warming impact category. Each box represents a unit process (only those >5% contribution to the impact are shown), with percentage contribution shown in bottom left, and quantity of the process for the assembly at the top of each box. The arrows represent the flow of materials between processes, and their thickness reflect the relative contribution.

**Supplementary Figure 4: Network diagram for cup fit FFP respirator showing global warming impact drivers (5% cut-off)**

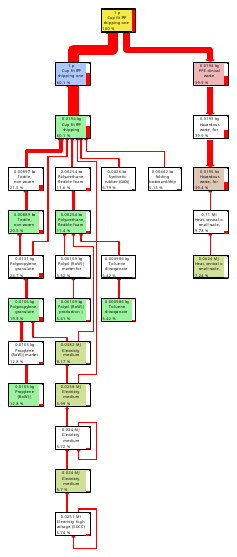

Network diagram for LCA of a single cup fit FFP respirator, showing drivers for the global warming impact category. Each box represents a unit process (only those >5% contribution to the impact are shown), with percentage contribution shown in bottom left, and quantity of the process for the assembly at the top of each box. The arrows represent the flow of materials between processes, and their thickness reflect the relative contribution. FFP= filtering facepiece

**Supplementary Figure 5: Network diagram for duckbill FFP respirator showing global warming impact drivers (5% cut-off)**

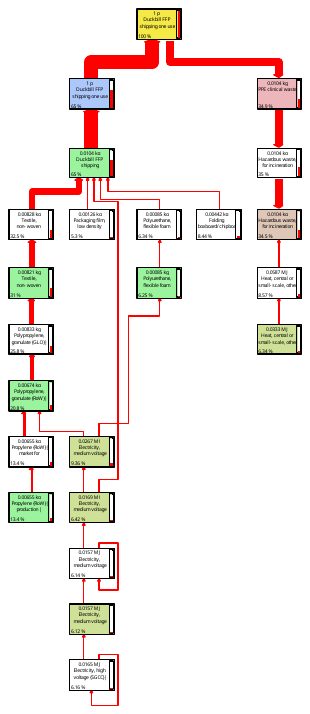

Network diagram for LCA of a single duckbill FFP respirator, showing drivers for the global warming impact category. Each box represents a unit process (only those >5% contribution to the impact are shown), with percentage contribution shown in bottom left, and quantity of the process for the assembly at the top of each box. The arrows represent the flow of materials between processes, and their thickness reflect the relative contribution. FFP= filtering facepiece

**Supplementary Figure 6: Network diagram for glove showing global warming impact drivers (5% cut-off)**

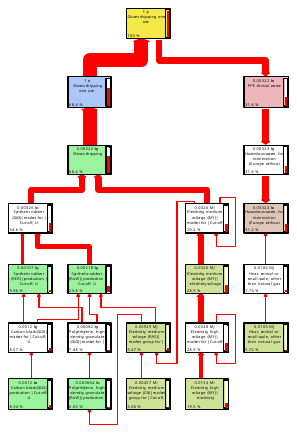

Network diagram for LCA of a single glove, showing drivers for the global warming impact category. Each box represents a unit process (only those >5% contribution to the impact are shown), with percentage contribution shown in bottom left, and quantity of the process for the assembly at the top of each box. The arrows represent the flow of materials between processes, and their thickness reflect the relative contribution.

**Supplementary Figure 7: Network diagram for disposable gown showing global warming impact drivers (5% cut-off)**

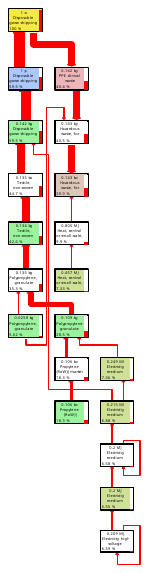

Network diagram for LCA of a single disposable gown, showing drivers for the global warming impact category. Each box represents a unit process (only those >5% contribution to the impact are shown), with percentage contribution shown in bottom left, and quantity of the process for the assembly at the top of each box. The arrows represent the flow of materials between processes, and their thickness reflect the relative contribution.

**Supplementary Figure 8: Network diagram for surgical mask (Type II) showing global warming impact drivers (5% cut-off)**

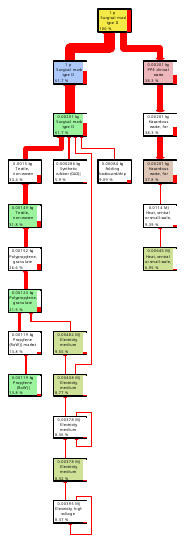

Network diagram for LCA of a single surgical mask (Type II), showing drivers for the global warming impact category. Each box represents a unit process (only those >5% contribution to the impact are shown), with percentage contribution shown in bottom left, and quantity of the process for the assembly at the top of each box. The arrows represent the flow of materials between processes, and their thickness reflect the relative contribution.

**Supplementary Figure 9: Network diagram for surgical mask (Type IIR) showing global warming impact drivers (5% cut-off)**

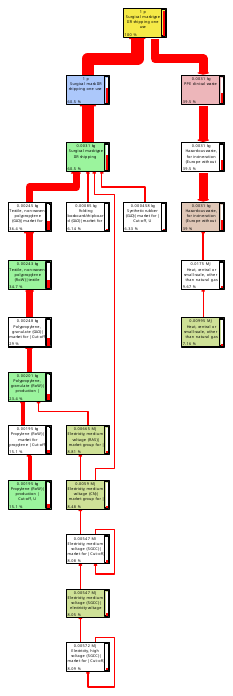

Network diagram for LCA of a single surgical mask (Type IIR), showing drivers for the global warming impact category. Each box represents a unit process (only those >5% contribution to the impact are shown), with percentage contribution shown in bottom left, and quantity of the process for the assembly at the top of each box. The arrows represent the flow of materials between processes, and their thickness reflect the relative contribution.

**Supplementary Figure 10: Comparison of environmental impacts of single-use versus reusable gowns**

Value compared to single-use gown (%)

|  | Single-use gown | Reusable gown |
| --- | --- | --- |
| Global warming | 100 | 32.6039 |
| Stratospheric ozone depletion | 100 | 44.2965 |
| Ionizing radiation | 100 | 82.8055 |
| Ozone formation, Human health | 100 | 25.6385 |
| Fine particulate matter formation | 100 | 29.649 |
| Ozone formation, Terrestrial ecosystems | 100 | 25.8652 |
| Terrestrial acidification | 100 | 26.7064 |
| Freshwater eutrophication | 100 | 23.7027 |
| Marine eutrophication | 67.9323 | 100 |
| Terrestrial ecotoxicity | 100 | 28.2665 |
| Freshwater ecotoxicity | 100 | 22.0138 |
| Marine ecotoxicity | 100 | 22.2466 |
| Human carcinogenic toxicity | 100 | 13.7845 |
| Human non-carcinogenic toxicity | 100 | 29.9891 |
| Land use | 46.2275 | 100 |
| Mineral resource scarcity | 100 | 30.0524 |
| Fossil resource scarcity | 100 | 30.4844 |
| Water consumption | 100 | 46.8415 |

Comparison of environmental impacts (midpoint categories) of single-use gown versus one use of reusable gown, normalised to highest scenario for each impact factor

**Supplementary Figure 11: Network diagram for a reusable gown showing marine eutrophication impact drivers (5% cut-off)**

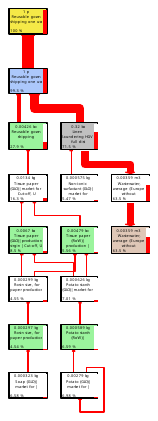

Network diagram for LCA of one use of a reusable gown showing drivers for the marine eutrophication impact category. Each box represents a unit process (only those >5% contribution to the impact are shown), with percentage contribution shown in bottom left, and quantity of the process for the assembly at the top of each box. The arrows represent the flow of materials between processes, and their thickness reflect the relative contribution.

**Supplementary figure 12: Network diagram for a reusable gown showing land use impact drivers (5% cut-off)**

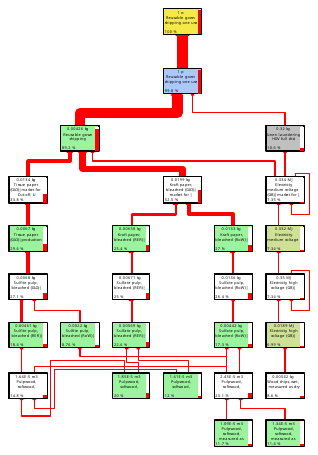

Network diagram for LCA of one use of a reusable gown showing drivers for the land use impact category. Each box represents a unit process (only those >5% contribution to the impact are shown), with percentage contribution shown in bottom left, and quantity of the process for the assembly at the top of each box. The arrows represent the flow of materials between processes, and their thickness reflect the relative contribution.

**Supplementary Figure 13: Comparison of environmental impacts of one-use versus re-use of face shields**

Value compared to single use of face shield (%)

|  | Single use of face shield | Reuse of face shield (x5) |
| --- | --- | --- |
| Global warming | 100 | 28.9146 |
| Stratospheric ozone depletion | 100 | 29.3688 |
| Ionizing radiation | 100 | 30.2514 |
| Ozone formation, Human health | 100 | 28.9173 |
| Fine particulate matter formation | 100 | 29.5241 |
| Ozone formation, Terrestrial ecosystems | 100 | 29.0208 |
| Terrestrial acidification | 100 | 29.2891 |
| Freshwater eutrophication | 100 | 30.0246 |
| Marine eutrophication | 100 | 29.0989 |
| Terrestrial ecotoxicity | 100 | 33.7268 |
| Freshwater ecotoxicity | 100 | 32.8131 |
| Marine ecotoxicity | 100 | 32.7298 |
| Human carcinogenic toxicity | 100 | 29.8305 |
| Human non-carcinogenic toxicity | 100 | 31.9657 |
| Land use | 100 | 32.117 |
| Mineral resource scarcity | 100 | 43.0945 |
| Fossil resource scarcity | 100 | 27.5244 |
| Water consumption | 100 | 26.574 |

Comparison of environmental impacts (midpoint categories) of single use of face shield, versus reuse of face shield five times with use of disinfectant wipe in between uses, normalised to highest scenario for each impact factor
