## Supplementary Figure 2 for "Environmental impact of Personal Protective Equipment distributed for use by health and social care services in England in the first six months of the COVID-19 pandemic"

**Supplementary table 1: transport assumptions**

| **Country of origin** | **Airport** | **Assumed distance for air freight to London Gatwick Airport (km)** | **Port** | **Assumed distance for shipping to London Gateway Port (km)** |
| --- | --- | --- | --- | --- |
| China | Beijing International | 7903 | Shanghai | 19303 |
| Egypt | Cairo International | 3503 | Damietta | 5952 |
| France | Charles de Gaulle | 310 | Le Harve | 347 |
| Germany | Frankfurt | 630 | Hamburg | 739 |
| Malaysia | Kuala Lumpa International | 10603 | Port Klang | 15026 |
| Mexico | Mexico City International | 8944 | Lázaro Cárdenas | 11868 |
| Thailand | Suvarnabhumi International | 9579 | Bangkok | 16950 |

Assumed airports, ports, and distances to the UK. We included 160km of travel by road via heavy goods vehicles both within the country of origin and in the UK, with an additional 8km either end of each journey via courier.

**Supplementary table 2: Life cycle inventory processes chosen for PPE**

| **Material** | **Process Name** |
| --- | --- |
| Aluminium | Sheet rolling, aluminium {GLO}\| market for \| Cut-off, U |
| Anionic surfactant | Anionic resin {RoW}\| market for anionic resin \| Cut-off, U |
| Box board | Folding boxboard/chipboard {GLO}\| market for \| Cut-off, U |
| Corrugated cardboard | Corrugated board box {RoW}\| market for corrugated board box \| Cut-off, U |
| LDPE film | Packaging film, low density polyethylene {GLO}\| market for \| Cut-off, U |
| Melt-blown polypropylene | Textile, non-woven polypropylene {GLO}\| market for textile, non woven polypropylene \| Cut-off, U |
| Non-woven polypropylene |  |
| **Nonionic surfactant** | Non-ionic surfactant {GLO}\| market for non-ionic surfactant \| Cut-off, U |
| Paper | Kraft paper, bleached {GLO}\| market for \| Cut-off, U |
| Polyester microfibre | Textile, non-woven polyester {GLO}\| market for textile, non woven polyester \| Cut-off, U |
| Polypropylene, injection moulded | Polypropylene, granulate {GLO}\| market for \| Cut-off, U |
| Polypropylene, oriented film |  |
| Polyurethane foam | Polyurethane, flexible foam {RoW}\| market for polyurethane, flexible foam \| Cut-off, U |
| Quaternary ammonium compounds and polymeric biguanide hydrochloride | Ammonia, liquid {RoW}\| market for \| Cut-off, U |
| Sodium hydroxide | Sodium hydroxide, without water, in 50% solution state {GLO}\| market for \| Cut-off, U |
| Sodium hypochlorite | Sodium hypochlorite, without water, in 15% solution state {RoW}\| market for sodium hypochlorite, without water, in 15% solution state \| Cut-off, U |
| Steel | Wire drawing, steel {GLO}\| market for \| Cut-off, U |
| Synthetic rubber | Synthetic rubber {GLO}\| market for \| Cut-off, U |
| Tissue paper | Tissue paper {GLO}\| market for \| Cut-off, U |
| **Electricity** | **Process Name** |
| Electricity (China) | Electricity, medium voltage {CN} market group for \| Cut-off, U |
| Electricity (France) | Electricity, medium voltage {FR}\| market for \| Cut-off, U |
| Electricity (UK) | Electricity, medium voltage {GB}\| market for \| Cut-off, U |
| Electricity (Malaysian) | Electricity, medium voltage {MY}\| market for \| Cut-off, U |
| **Transportation** | **Process Name** |
| Air freight | Transport, freight, aircraft, long haul {GLO}\| market for transport, freight, aircraft, long haul \| Cut-off, U |
| Courier | Transport, freight, light commercial vehicle {GLO}\| market group for transport, freight, light commercial vehicle \| Cut-off, U |
| Heavy goods vehicle | Transport, freight, lorry, unspecified {GLO}\| market group for transport, freight, lorry, unspecified \| Cut-off, U |
| Shipping container | Transport, freight, sea, container ship {GLO}\| market for transport, freight, sea, container ship \| Cut-off, U |
| **Waste** | **Process Name** |
| Hazardous incineration | Hazardous waste, for incineration {Europe without Switzerland}\| treatment of hazardous waste, hazardous waste incineration \| Cut-off, U |

Processes selected for PPE LCA. Ecoinvent (version 3.6)- allocation, cut-off by classification- unit library was selected for all processes.

**Supplementary table 3: linen laundering assumptions**

| ****Input**** | | | ****Source**** | ****Quantity used per kg of dry linen**** |
| --- | --- | --- | --- | --- |
| **Detergent** | | **Anionic surfactant**  **Nonionic surfactant**  **Sodium hydroxide**  **Sodium hypochlorite** | Carre^1^ | **0.54 g**  **1.8 g**  **1.92 g**  **1.1 g** |
| **Electricity** | | | Vozzola et al.^2^ | **0.28 kWh** |
| **Natural gas** | | |  | **1.59 kWh** |
| **Transport via heavy goods vehicle** | | | Assumed | **160 kg.km** |
| **Water** | **Supply** | | Vozzola et al.^2^ | **0.01100** m^3^ |
|  | **Treatment** | |  | **0.01097** m^3^ |

kWh= kilowatt hours, m^3^= meters cubed, kg.km= kilogram kilometres.

1. Carre A. Life cycle assessment comparing laundered surgical gowns with polypropylene based disposable gowns. Melbourne, Australia: RMIT University; 2008.

2. Vozzola E, Overcash M, Griffing E. Environmental considerations in the selection of isolation gowns: A life cycle assessment of reusable and disposable alternatives. Am J Infect Control. 2018;46(8):881-6.

**Supplementary table 4: Environmental impact of one apron**

| **Impact category** | **Unit** | **Total** | Packaging film, low density polyethylene {GLO}\| market for \| Cut-off, U | Corrugated board box {RoW}\| market for corrugated board box \| Cut-off, U | Packaging film, low density polyethylene {GLO}\| market for \| Cut-off, U | Transport, freight, sea, container ship {GLO}\| market for transport, freight, sea, container ship \| Cut-off, U | Transport, freight, light commercial vehicle {GLO}\| market group for transport, freight, light commercial vehicle \| Cut-off, U | Transport, freight, lorry, unspecified {GLO}\| market group for transport, freight, lorry, unspecified \| Cut-off, U | Electricity, medium voltage {CN}\| market group for \| Cut-off, U | Electricity, medium voltage {TH}\| market for \| Cut-off, U | PPE clinical waste |
| --- | --- | --- | --- | --- | --- | --- | --- | --- | --- | --- | --- |
| **Global warming** | kg CO_2_e | 0.065280133 | 0.031777301 | 0.000245003 | 0.000102487 | 0.0018351 | 0.000608815 | 0.000441633 | 0.004535052 | 0.000390236 | 0.02534449 |
| **Stratospheric ozone depletion** | kg CFC11 eq | 2.03E-08 | 8.06196E-09 | 2.44109E-10 | 2.60011E-11 | 1.29E-09 | 3.67556E-10 | 2.06736E-10 | 9.66461E-10 | 1.15915E-10 | 9.01E-09 |
| **Ionizing radiation** | kBq Co-60 eq | 0.00251773 | 0.002001859 | 7.30242E-06 | 6.4563E-06 | 1.91E-05 | 1.88829E-05 | 8.62711E-06 | 6.95485E-05 | 8.5941E-07 | 0.000385128 |
| **Ozone formation, Human health** | kg NO_x_ eq | 0.000156504 | 7.67665E-05 | 7.65766E-07 | 2.47584E-07 | 3.82E-05 | 2.7308E-06 | 2.39737E-06 | 1.25632E-05 | 5.3224E-07 | 2.23E-05 |
| **Fine particulate matter formation** | kg PM2.5 eq | 8.14E-05 | 4.2836E-05 | 4.02437E-07 | 1.38153E-07 | 1.22E-05 | 9.71347E-07 | 6.43614E-07 | 6.92792E-06 | 3.16651E-07 | 1.70E-05 |
| **Ozone formation, Terrestrial ecosystems** | kg NOx eq | 0.000163703 | 8.30787E-05 | 7.81684E-07 | 2.67941E-07 | 3.84E-05 | 2.81687E-06 | 2.44532E-06 | 1.25898E-05 | 5.42528E-07 | 2.27E-05 |
| **Terrestrial acidification** | kg SO_2_ eq | 0.000196905 | 9.40803E-05 | 8.65169E-07 | 3.03423E-07 | 3.79E-05 | 2.11228E-06 | 1.48405E-06 | 1.54635E-05 | 8.33185E-07 | 4.39E-05 |
| **Freshwater eutrophication** | kg P eq | 1.91E-05 | 8.88645E-06 | 9.74318E-08 | 2.86602E-08 | 7.22E-08 | 1.12707E-07 | 4.05883E-08 | 8.27533E-07 | 2.40319E-07 | 8.84E-06 |
| **Marine eutrophication** | kg N eq | 1.75E-06 | 1.08866E-06 | 5.53837E-08 | 3.5111E-09 | 6.93E-09 | 6.79274E-09 | 3.2385E-09 | 5.35525E-08 | 1.57581E-08 | 5.12E-07 |
| **Terrestrial ecotoxicity** | kg 1,4-DCB | 0.1159339 | 0.065167219 | 0.001024047 | 0.000210174 | 0.004892366 | 0.00400804 | 0.008440728 | 0.002995972 | 0.000201959 | 0.028993398 |
| **Freshwater ecotoxicity** | kg 1,4-DCB | 0.001721327 | 0.001011849 | 1.08516E-05 | 3.26337E-06 | 1.97E-05 | 5.91267E-05 | 1.03475E-05 | 6.96628E-05 | 1.01443E-05 | 0.000526426 |
| **Marine ecotoxicity** | kg 1,4-DCB | 0.002293897 | 0.001332203 | 1.42195E-05 | 4.29656E-06 | 2.81E-05 | 7.82752E-05 | 1.80408E-05 | 9.21552E-05 | 1.35003E-05 | 0.000713144 |
| **Human carcinogenic toxicity** | kg 1,4-DCB | 0.003935434 | 0.000986104 | 7.26716E-06 | 3.18034E-06 | 4.14E-05 | 2.72105E-05 | 1.01608E-05 | 0.000127373 | 1.41123E-05 | 0.002718647 |
| **Human non-carcinogenic toxicity** | kg 1,4-DCB | 0.039912556 | 0.021923231 | 0.000264608 | 7.07058E-05 | 0.000327085 | 0.00119536 | 0.000340164 | 0.002145158 | 0.000295157 | 0.013351086 |
| **Land use** | m^2^a crop eq | 0.001441517 | 0.001038614 | 0.000109955 | 3.34969E-06 | 6.52E-06 | 1.64576E-05 | 2.41682E-05 | 5.43358E-05 | 1.37524E-06 | 1.87E-04 |
| **Mineral resource scarcity** | kg Cu eq | 0.000100139 | 6.58362E-05 | 7.16812E-07 | 2.12332E-07 | 4.63E-06 | 2.90321E-06 | 1.65254E-06 | 1.88885E-06 | 2.70822E-07 | 2.20E-05 |
| **Fossil resource scarcity** | kg oil eq | 0.022970775 | 0.018777169 | 6.9035E-05 | 6.05592E-05 | 0.00054157 | 0.000198108 | 0.000152517 | 0.000880587 | 0.000114521 | 0.002176709 |
| **Water consumption** | m^3^ | 0.000648351 | 0.000541592 | 2.92374E-06 | 1.74672E-06 | 1.22E-06 | 1.49607E-06 | 8.13631E-07 | 1.11521E-05 | 1.57455E-06 | 8.58E-05 |

Environmental impacts (midpoint categories) measured using life cycle assessment modelled on one apron (with shipping, and waste via hazardous incineration). 1,4-DCB =dichlorobenzene, CFC11= Trichlorofluoromethane, CO_2_e= carbon dioxide equivalents, Cu= copper, eq= equivalents, kBq Co-60 eq = kilobecquerel Cobalt-60, m^2^a = square meter years, N= nitrogen, NO_x_= nitrous oxides, P=phosphate, PM2.5 = particulate matter <2.5 micrometers, SO_2_= sulphur dioxide

**Supplementary Table 5: Environmental impact of one face shield**

| **Impact category** | **Unit** | **Total** | Packaging film, low density polyethylene {GLO}\| market for \| Cut-off, U | Polyurethane, flexible foam {RoW}\| market for polyurethane, flexible foam \| Cut-off, U | Synthetic rubber {GLO}\| market for \| Cut-off, U | Packaging film, low density polyethylene {GLO}\| market for \| Cut-off, U | Corrugated board box {RoW}\| market for corrugated board box \| Cut-off, U | Transport, freight, lorry, unspecified {GLO}\| market group for transport, freight, lorry, unspecified \| Cut-off, U | Transport, freight, light commercial vehicle {GLO}\| market group for transport, freight, light commercial vehicle \| Cut-off, U | Transport, freight, sea, container ship {GLO}\| market for transport, freight, sea, container ship \| Cut-off, U | Electricity, medium voltage {CN}\| market group for \| Cut-off, U | PPE clinical waste |
| --- | --- | --- | --- | --- | --- | --- | --- | --- | --- | --- | --- | --- |
| **Global warming** | kg CO_2_e | 0.231369 | 0.059719 | 0.030962 | 0.013935 | 0.02733 | 0.002291 | 0.001757 | 0.002422 | 0.007402 | 0.011212 | 0.074337 |
| **Stratospheric ozone depletion** | kg CFC11 eq | 7.22E-08 | 1.52E-08 | 2.66E-09 | 8.88E-09 | 6.93E-09 | 2.28E-09 | 8.23E-10 | 1.46E-09 | 5.19E-09 | 2.39E-09 | 2.64E-08 |
| **Ionizing radiation** | kBq Co-60 eq | 0.008239 | 0.003762 | 0.000101 | 0.001098 | 0.001722 | 6.83E-05 | 3.43E-05 | 7.51E-05 | 7.69E-05 | 0.000172 | 0.00113 |
| **Ozone formation, Human health** | kg NO_x_ eq | 0.00059 | 0.000144 | 6.54E-05 | 3.64E-05 | 6.60E-05 | 7.16E-06 | 9.54E-06 | 1.09E-05 | 0.000154 | 3.11E-05 | 6.55E-05 |
| **Fine particulate matter formation** | kg PM2.5 eq | 0.000309 | 8.05E-05 | 4.03E-05 | 2.48E-05 | 3.68E-05 | 3.76E-06 | 2.56E-06 | 3.86E-06 | 4.92E-05 | 1.71E-05 | 4.97E-05 |
| **Ozone formation, Terrestrial ecosystems** | kg NOx eq | 0.000616 | 0.000156 | 6.82E-05 | 3.95E-05 | 7.15E-05 | 7.31E-06 | 9.73E-06 | 1.12E-05 | 0.000155 | 3.11E-05 | 6.67E-05 |
| **Terrestrial acidification** | kg SO_2_ eq | 0.000757 | 0.000177 | 0.000103 | 5.39E-05 | 8.09E-05 | 8.09E-06 | 5.90E-06 | 8.40E-06 | 0.000153 | 3.82E-05 | 1.29E-04 |
| **Freshwater eutrophication** | kg P eq | 6.16E-05 | 1.67E-05 | 3.06E-06 | 4.39E-06 | 7.64E-06 | 9.11E-07 | 1.61E-07 | 4.48E-07 | 2.91E-07 | 2.05E-06 | 2.59E-05 |
| **Marine eutrophication** | kg N eq | 1.12E-05 | 2.05E-06 | 5.71E-06 | 3.18E-07 | 9.36E-07 | 5.18E-07 | 1.29E-08 | 2.70E-08 | 2.80E-08 | 1.32E-07 | 1.50E-06 |
| **Terrestrial ecotoxicity** | kg 1,4-DCB | 0.431942 | 0.122469 | 0.030581 | 0.051558 | 0.056046 | 0.009578 | 0.033584 | 0.015947 | 0.019733 | 0.007407 | 0.085039 |
| **Freshwater ecotoxicity** | kg 1,4-DCB | 0.006321 | 0.001902 | 0.000513 | 0.000863 | 0.00087 | 0.000101 | 4.12E-05 | 0.000235 | 7.93E-05 | 0.000172 | 0.001544 |
| **Marine ecotoxicity** | kg 1,4-DCB | 0.008418 | 0.002504 | 0.000678 | 0.001142 | 0.001146 | 0.000133 | 7.18E-05 | 0.000311 | 0.000113 | 0.000228 | 0.002092 |
| **Human carcinogenic toxicity** | kg 1,4-DCB | 0.012861 | 0.001853 | 0.000937 | 0.000551 | 0.000848 | 6.80E-05 | 4.04E-05 | 0.000108 | 0.000167 | 0.000315 | 0.007974 |
| **Human non-carcinogenic toxicity** | kg 1,4-DCB | 0.14318 | 0.041201 | 0.009599 | 0.019159 | 0.018855 | 0.002475 | 0.001353 | 0.004756 | 0.001319 | 0.005304 | 0.03916 |
| **Land use** | m^2^a crop eq | 0.005291 | 0.001952 | 8.99E-05 | 0.000457 | 0.000893 | 0.001028 | 9.62E-05 | 6.55E-05 | 2.63E-05 | 0.000134 | 0.000548 |
| **Mineral resource scarcity** | kg Cu eq | 0.000738 | 0.000124 | 2.56E-05 | 0.000419 | 5.66E-05 | 6.70E-06 | 6.58E-06 | 1.16E-05 | 1.87E-05 | 4.67E-06 | 6.46E-05 |
| **Fossil resource scarcity** | kg oil eq | 0.083638 | 0.035288 | 0.01082 | 0.008594 | 0.016149 | 0.000646 | 0.000607 | 0.000788 | 0.002184 | 0.002177 | 0.006384 |
| **Water consumption** | m^3^ | 0.002692 | 0.001018 | 0.000662 | 0.000225 | 0.000466 | 2.73E-05 | 3.24E-06 | 5.95E-06 | 4.93E-06 | 2.76E-05 | 2.52E-04 |

Environmental impacts (midpoint categories) measured using life cycle assessment modelled on one face shield (with shipping, and waste via hazardous incineration). 1,4-DCB =dichlorobenzene, CFC11= Trichlorofluoromethane, CO_2_e= carbon dioxide equivalents, Cu= copper, eq= equivalents, kBq Co-60 eq = kilobecquerel Cobalt-60, m^2^a = square meter years, N= nitrogen, NO_x_= nitrous oxides, P=phosphate, PM2.5 = particulate matter <2.5 micrometers, SO_2_= sulphur dioxide

**Supplementary Table 6: Environmental impact of one cup fit FFP respirator**

| **Impact category** | **Unit** | **Total** | Textile, non-woven polypropylene {GLO}\| market for textile, non woven polypropylene \| Cut-off, U | Polypropylene, granulate {GLO}\| market for \| Cut-off, U | Polyurethane, flexible foam {RoW}\| market for polyurethane, flexible foam \| Cut-off, U | Synthetic rubber {GLO}\| market for \| Cut-off, U | Sheet rolling, aluminium {GLO}\| market for \| Cut-off, U | Wire drawing, steel {GLO}\| market for \| Cut-off, U | Corrugated board box {RoW}\| market for corrugated board box \| Cut-off, U | Folding boxboard/chipboard {GLO}\| market for \| Cut-off, U | Transport, freight, sea, container ship {GLO}\| market for transport, freight, sea, container ship \| Cut-off, U | Transport, freight, light commercial vehicle {GLO}\| market group for transport, freight, light commercial vehicle \| Cut-off, U | Transport, freight, lorry, unspecified {GLO}\| market group for transport, freight, lorry, unspecified \| Cut-off, U | Electricity, medium voltage {FR}\| market for \| Cut-off, U | Electricity, medium voltage {GB}\| market for \| Cut-off, U | Electricity, medium voltage {CN}\| market group for \| Cut-off, U | PPE clinical waste |
| --- | --- | --- | --- | --- | --- | --- | --- | --- | --- | --- | --- | --- | --- | --- | --- | --- | --- |
| **Global warming** | kg CO_2_e | 0.125163 | 0.026924 | 0.008633 | 0.014457 | 0.007179 | 0.000741 | 0.000109 | 0.001956 | 0.00645 | 0.002428 | 0.001589 | 0.001153 | 0.000167 | 0.000722 | 0.002023 | 0.049969 |
| **Stratospheric ozone depletion** | kg CFC11 eq | 4.26E-08 | 7.69E-09 | 1.54E-09 | 1.24E-09 | 4.58E-09 | 2.58E-10 | 3.96E-11 | 1.95E-09 | 3.21E-09 | 1.70E-09 | 9.60E-10 | 5.40E-10 | 2.31E-10 | 3.71E-10 | 4.31E-10 | 1.78E-08 |
| **Ionizing radiation** | kBq Co-60 eq | 0.005289 | 0.001537 | 0.000173 | 4.71E-05 | 0.000566 | 5.52E-05 | 3.88E-06 | 5.83E-05 | 0.000453 | 2.52E-05 | 4.93E-05 | 2.25E-05 | 0.00108 | 4.16E-04 | 3.10E-05 | 0.000759 |
| **Ozone formation, Human health** | kg NO_x_ eq | 0.000273 | 6.38E-05 | 1.78E-05 | 3.05E-05 | 1.88E-05 | 1.68E-06 | 1.95E-07 | 6.11E-06 | 1.75E-05 | 5.05E-05 | 7.13E-06 | 6.26E-06 | 3.08E-07 | 1.26E-06 | 5.60E-06 | 4.40E-05 |
| **Fine particulate matter formation** | kg PM2.5 eq | 0.000152 | 3.58E-05 | 9.03E-06 | 1.88E-05 | 1.28E-05 | 1.41E-06 | 1.67E-07 | 3.21E-06 | 1.23E-05 | 1.61E-05 | 2.54E-06 | 1.68E-06 | 1.94E-07 | 5.85E-07 | 3.09E-06 | 3.34E-05 |
| **Ozone formation, Terrestrial ecosystems** | kg NOx eq | 0.000282 | 6.69E-05 | 1.89E-05 | 3.18E-05 | 2.04E-05 | 1.76E-06 | 2.04E-07 | 6.24E-06 | 1.78E-05 | 5.09E-05 | 7.35E-06 | 6.38E-06 | 3.14E-07 | 1.27E-06 | 5.62E-06 | 4.48E-05 |
| **Terrestrial acidification** | kg SO_2_ eq | 0.000372 | 8.07E-05 | 2.34E-05 | 4.80E-05 | 2.78E-05 | 2.48E-06 | 2.88E-07 | 6.91E-06 | 2.59E-05 | 5.01E-05 | 5.51E-06 | 3.87E-06 | 4.63E-07 | 1.70E-06 | 6.90E-06 | 8.65E-05 |
| **Freshwater eutrophication** | kg P eq | 3.45E-05 | 7.24E-06 | 1.54E-06 | 1.43E-06 | 2.26E-06 | 2.90E-07 | 5.77E-08 | 7.78E-07 | 2.26E-06 | 9.55E-08 | 2.94E-07 | 1.06E-07 | 4.99E-08 | 1.52E-07 | 3.69E-07 | 1.74E-05 |
| **Marine eutrophication** | kg N eq | 5.64E-06 | 6.55E-07 | 1.27E-07 | 2.67E-06 | 1.64E-07 | 2.44E-08 | 8.03E-09 | 4.42E-07 | 4.34E-07 | 9.17E-09 | 1.77E-08 | 8.45E-09 | 2.26E-08 | 1.71E-08 | 2.39E-08 | 1.01E-06 |
| **Terrestrial ecotoxicity** | kg 1,4-DCB | 0.246403 | 0.062767 | 0.016589 | 0.014279 | 0.02656 | 0.000851 | 0.000197 | 0.008177 | 0.016734 | 0.006473 | 0.010463 | 0.022035 | 0.000719 | 0.000786 | 0.001336 | 0.057163 |
| **Freshwater ecotoxicity** | kg 1,4-DCB | 0.003614 | 0.00105 | 0.000253 | 0.000239 | 0.000445 | 3.35E-05 | 5.57E-06 | 8.67E-05 | 0.000172 | 2.60E-05 | 0.000154 | 2.70E-05 | 1.47E-05 | 1.91E-05 | 3.11E-05 | 0.001038 |
| **Marine ecotoxicity** | kg 1,4-DCB | 0.004795 | 0.001362 | 0.00033 | 0.000316 | 0.000588 | 4.33E-05 | 7.49E-06 | 0.000114 | 0.000229 | 3.71E-05 | 0.000204 | 4.71E-05 | 1.86E-05 | 2.60E-05 | 4.11E-05 | 0.001406 |
| **Human carcinogenic toxicity** | kg 1,4-DCB | 0.007786 | 0.000835 | 0.000226 | 0.000437 | 0.000284 | 5.48E-05 | 8.37E-05 | 5.80E-05 | 0.000196 | 5.48E-05 | 7.10E-05 | 2.65E-05 | 7.94E-06 | 1.70E-05 | 5.68E-05 | 0.00536 |
| **Human non-carcinogenic toxicity** | kg 1,4-DCB | 0.078678 | 0.018525 | 0.00476 | 0.004482 | 0.00987 | 0.000636 | 0.000133 | 0.002113 | 0.005382 | 0.000433 | 0.00312 | 0.000888 | 0.00026 | 0.00043 | 0.000957 | 0.026323 |
| **Land use** | m^2^a crop eq | 0.004457 | 0.000319 | 6.93E-05 | 4.20E-05 | 0.000236 | 9.44E-06 | 4.17E-06 | 0.000878 | 0.0023 | 8.63E-06 | 4.30E-05 | 6.31E-05 | 7.47E-06 | 8.00E-05 | 2.42E-05 | 0.000368 |
| **Mineral resource scarcity** | kg Cu eq | 0.000414 | 7.23E-05 | 2.11E-05 | 1.20E-05 | 0.000216 | 2.80E-06 | 2.47E-06 | 5.72E-06 | 1.50E-05 | 6.13E-06 | 7.58E-06 | 4.31E-06 | 1.40E-06 | 9.11E-07 | 8.42E-07 | 4.34E-05 |
| **Fossil resource scarcity** | kg oil eq | 0.042231 | 0.016966 | 0.006246 | 0.005052 | 0.004427 | 0.00019 | 2.03E-05 | 0.000551 | 0.001693 | 0.000717 | 0.000517 | 0.000398 | 4.65E-05 | 0.000244 | 0.000393 | 0.004292 |
| **Water consumption** | m^3^ | 0.00107 | 0.000257 | 7.53E-05 | 0.000309 | 0.000116 | 5.75E-06 | 5.17E-06 | 2.33E-05 | 8.05E-05 | 1.62E-06 | 3.91E-06 | 2.12E-06 | 6.38E-06 | 3.16E-06 | 4.97E-06 | 1.69E-04 |

Environmental impacts (midpoint categories) measured using life cycle assessment modelled on one cup fit respirator (with shipping, and waste via hazardous incineration). 1,4-DCB =dichlorobenzene, CFC11= Trichlorofluoromethane, CO_2_e= carbon dioxide equivalents, Cu= copper, eq= equivalents, FFP= filtering facepiece, kBq Co-60 eq = kilobecquerel Cobalt-60, m^2^a = square meter years, N= nitrogen, NO_x_= nitrous oxides, P=phosphate, PM2.5 = particulate matter <2.5 micrometers, SO_2_= sulphur dioxide

**Supplementary Table 7: Environmental impact of one duckbill FFP respirator**

| **Impact category** | **Unit** | **Total** | Textile, non-woven polypropylene {GLO}\| market for textile, non woven polypropylene \| Cut-off, U | Synthetic rubber {GLO}\| market for \| Cut-off, U | Polyurethane, flexible foam {RoW}\| market for polyurethane, flexible foam \| Cut-off, U | Wire drawing, steel {GLO}\| market for \| Cut-off, U | Corrugated board box {RoW}\| market for corrugated board box \| Cut-off, U | Folding boxboard/chipboard {GLO}\| market for \| Cut-off, U | Packaging film, low density polyethylene {GLO}\| market for \| Cut-off, U | Transport, freight, sea, container ship {GLO}\| market for transport, freight, sea, container ship \| Cut-off, U | Transport, freight, light commercial vehicle {GLO}\| market group for transport, freight, light commercial vehicle \| Cut-off, U | Transport, freight, lorry, unspecified {GLO}\| market group for transport, freight, lorry, unspecified \| Cut-off, U | Electricity, medium voltage {CN}\| market group for \| Cut-off, U | Electricity, medium voltage {FR}\| market for \| Cut-off, U | Electricity, medium voltage {GB}\| market for \| Cut-off, U | PPE clinical waste |
| --- | --- | --- | --- | --- | --- | --- | --- | --- | --- | --- | --- | --- | --- | --- | --- | --- |
| **Global warming** | kg CO_2_e | 0.07635 | 0.024853 | 0.002168 | 0.004838 | 0.000163 | 0.001956 | 0.006445 | 0.004029 | 0.00172 | 0.001126 | 0.000817 | 1.08E-03 | 8.93E-05 | 0.000386 | 0.026681 |
| **Stratospheric ozone depletion** | kg CFC11 eq | 2.74E-08 | 7.10E-09 | 1.38E-09 | 4.15E-10 | 5.94E-11 | 1.95E-09 | 3.21E-09 | 1.02E-09 | 1.21E-09 | 6.80E-10 | 3.82E-10 | 2.30E-10 | 1.23E-10 | 1.98E-10 | 9.48E-09 |
| **Ionizing radiation** | kBq Co-60 eq | 0.003665 | 0.001419 | 0.000171 | 1.58E-05 | 5.82E-06 | 5.83E-05 | 0.000452 | 2.54E-04 | 1.79E-05 | 3.49E-05 | 1.60E-05 | 1.66E-05 | 0.000577 | 0.000222 | 0.000405 |
| **Ozone formation, Human health** | kg NO_x_ eq | 0.000181 | 5.89E-05 | 5.66E-06 | 1.02E-05 | 2.93E-07 | 6.11E-06 | 1.75E-05 | 9.73E-06 | 3.58E-05 | 5.05E-06 | 4.43E-06 | 2.99E-06 | 1.64E-07 | 6.74E-07 | 2.35E-05 |
| **Fine particulate matter formation** | kg PM2.5 eq | 9.88E-05 | 3.31E-05 | 3.86E-06 | 6.29E-06 | 2.50E-07 | 3.21E-06 | 1.23E-05 | 5.43E-06 | 1.14E-05 | 1.80E-06 | 1.19E-06 | 1.65E-06 | 1.03E-07 | 3.13E-07 | 1.78E-05 |
| **Ozone formation, Terrestrial ecosystems** | kg NOx eq | 0.000187 | 6.18E-05 | 6.14E-06 | 1.07E-05 | 3.06E-07 | 6.24E-06 | 1.77E-05 | 1.05E-05 | 3.60E-05 | 5.21E-06 | 4.52E-06 | 3.00E-06 | 1.67E-07 | 6.81E-07 | 2.39E-05 |
| **Terrestrial acidification** | kg SO_2_ eq | 0.000237 | 7.45E-05 | 8.38E-06 | 1.61E-05 | 4.33E-07 | 6.91E-06 | 2.58E-05 | 1.19E-05 | 3.55E-05 | 3.91E-06 | 2.74E-06 | 3.68E-06 | 2.47E-07 | 9.09E-07 | 4.62E-05 |
| **Freshwater eutrophication** | kg P eq | 2.21E-05 | 6.68E-06 | 6.83E-07 | 4.78E-07 | 8.65E-08 | 7.78E-07 | 2.26E-06 | 1.13E-06 | 6.76E-08 | 2.08E-07 | 7.51E-08 | 1.97E-07 | 2.66E-08 | 8.11E-08 | 9.31E-06 |
| **Marine eutrophication** | kg N eq | 3.17E-06 | 6.05E-07 | 4.95E-08 | 8.93E-07 | 1.21E-08 | 4.42E-07 | 4.33E-07 | 1.38E-07 | 6.50E-09 | 1.26E-08 | 5.99E-09 | 1.28E-08 | 1.21E-08 | 9.14E-09 | 5.39E-07 |
| **Terrestrial ecotoxicity** | kg 1,4-DCB | 0.16384 | 0.057939 | 0.00802 | 0.004778 | 0.000296 | 0.008177 | 0.016719 | 0.008262 | 0.004586 | 0.007413 | 0.015611 | 0.000713 | 0.000384 | 0.00042 | 0.030522 |
| **Freshwater ecotoxicity** | kg 1,4-DCB | 0.002314 | 0.000969 | 0.000134 | 8.01E-05 | 8.36E-06 | 8.67E-05 | 0.000171 | 1.28E-04 | 1.84E-05 | 0.000109 | 1.91E-05 | 1.66E-05 | 7.84E-06 | 1.02E-05 | 0.000554 |
| **Marine ecotoxicity** | kg 1,4-DCB | 0.003064 | 0.001257 | 0.000178 | 0.000106 | 1.12E-05 | 0.000114 | 0.000229 | 0.000169 | 2.63E-05 | 0.000145 | 3.34E-05 | 2.19E-05 | 9.94E-06 | 1.39E-05 | 0.000751 |
| **Human carcinogenic toxicity** | kg 1,4-DCB | 0.004521 | 0.00077 | 8.56E-05 | 0.000146 | 1.26E-04 | 5.80E-05 | 0.000196 | 0.000125 | 3.88E-05 | 5.03E-05 | 1.88E-05 | 3.03E-05 | 4.24E-06 | 9.10E-06 | 0.002862 |
| **Human non-carcinogenic toxicity** | kg 1,4-DCB | 0.05013 | 0.0171 | 0.00298 | 0.0015 | 0.0002 | 0.002113 | 0.005377 | 0.002779 | 0.000307 | 0.002211 | 0.000629 | 0.000511 | 0.000139 | 0.00023 | 0.014055 |
| **Land use** | m^2^a crop eq | 0.004031 | 0.000294 | 7.11E-05 | 1.40E-05 | 6.25E-06 | 0.000878 | 0.002298 | 1.32E-04 | 6.12E-06 | 3.04E-05 | 4.47E-05 | 1.29E-05 | 3.99E-06 | 4.27E-05 | 1.97E-04 |
| **Mineral resource scarcity** | kg Cu eq | 0.000206 | 6.67E-05 | 6.52E-05 | 4.00E-06 | 3.70E-06 | 5.72E-06 | 1.50E-05 | 8.35E-06 | 4.34E-06 | 5.37E-06 | 3.06E-06 | 4.50E-07 | 7.46E-07 | 4.86E-07 | 2.32E-05 |
| **Fossil resource scarcity** | kg oil eq | 0.027154 | 0.015661 | 0.001337 | 0.001691 | 3.05E-05 | 0.000551 | 0.001691 | 0.00238 | 0.000508 | 0.000366 | 0.000282 | 2.10E-04 | 2.48E-05 | 0.00013 | 0.002292 |
| **Water consumption** | m^3^ | 0.00066 | 0.000237 | 3.51E-05 | 0.000104 | 7.76E-06 | 2.33E-05 | 8.04E-05 | 6.87E-05 | 1.15E-06 | 2.77E-06 | 1.50E-06 | 2.66E-06 | 3.41E-06 | 1.69E-06 | 9.04E-05 |

Environmental impacts (midpoint categories) measured using life cycle assessment modelled on one duckbill FFP respirator (with shipping, and waste via hazardous incineration). 1,4-DCB =dichlorobenzene, CFC11= Trichlorofluoromethane, CO_2_e= carbon dioxide equivalents, Cu= copper, eq= equivalents, FFP=filtering facepiece, kBq Co-60 eq = kilobecquerel Cobalt-60, m^2^a = square meter years, N= nitrogen, NO_x_= nitrous oxides, P=phosphate, PM2.5 = particulate matter <2.5 micrometers, SO_2_= sulphur dioxide

**Supplementary Table 8: Environmental impact of one glove**

| **Impact category** | **Unit** | **Total** | Synthetic rubber {GLO}\| market for \| Cut-off, U | Corrugated board box {RoW}\| market for corrugated board box \| Cut-off, U | Folding boxboard/chipboard {GLO}\| market for \| Cut-off, U | Transport, freight, lorry, unspecified {GLO}\| market group for transport, freight, lorry, unspecified \| Cut-off, U | Transport, freight, light commercial vehicle {GLO}\| market group for transport, freight, light commercial vehicle \| Cut-off, U | Transport, freight, sea, container ship {GLO}\| market for transport, freight, sea, container ship \| Cut-off, U | Electricity, medium voltage {MY}\| market for \| Cut-off, U | PPE clinical waste |
| --- | --- | --- | --- | --- | --- | --- | --- | --- | --- | --- |
| **Global warming** | kg CO_2_e | 0.02624 | 0.009079 | 0.000127 | 0.000234 | 0.000154 | 0.000212 | 0.000505 | 0.00764 | 0.00829 |
| **Stratospheric ozone depletion** | kg CFC11 eq | 1.17E-08 | 5.79E-09 | 1.27E-10 | 1.16E-10 | 7.21E-11 | 1.28E-10 | 3.54E-10 | 2.12E-09 | 2.95E-09 |
| **Ionizing radiation** | kBq Co-60 eq | 0.000889 | 0.000716 | 3.79E-06 | 1.64E-05 | 3.01E-06 | 6.58E-06 | 5.24E-06 | 1.24E-05 | 1.26E-04 |
| **Ozone formation, Human health** | kg NO_x_ eq | 5.63E-05 | 2.37E-05 | 3.98E-07 | 6.34E-07 | 8.36E-07 | 9.52E-07 | 1.05E-05 | 1.19E-05 | 7.31E-06 |
| **Fine particulate matter formation** | kg PM2.5 eq | 4.55E-05 | 1.62E-05 | 2.09E-07 | 4.47E-07 | 2.24E-07 | 3.39E-07 | 3.35E-06 | 1.92E-05 | 5.55E-06 |
| **Ozone formation, Terrestrial ecosystems** | kg NOx eq | 5.87E-05 | 2.57E-05 | 4.06E-07 | 6.43E-07 | 8.52E-07 | 9.82E-07 | 1.06E-05 | 1.21E-05 | 7.44E-06 |
| **Terrestrial acidification** | kg SO_2_ eq | 8.77E-05 | 3.51E-05 | 4.50E-07 | 9.36E-07 | 5.17E-07 | 7.36E-07 | 1.04E-05 | 2.51E-05 | 1.44E-05 |
| **Freshwater eutrophication** | kg P eq | 9.22E-06 | 2.86E-06 | 5.06E-08 | 8.17E-08 | 1.41E-08 | 3.93E-08 | 1.98E-08 | 3.26E-06 | 2.89E-06 |
| **Marine eutrophication** | kg N eq | 6.41E-07 | 2.07E-07 | 2.88E-08 | 1.57E-08 | 1.13E-09 | 2.37E-09 | 1.91E-09 | 2.17E-07 | 1.67E-07 |
| **Terrestrial ecotoxicity** | kg 1,4-DCB | 0.054753 | 0.033591 | 0.000532 | 0.000606 | 0.002942 | 0.001397 | 0.001346 | 0.004857 | 0.009483 |
| **Freshwater ecotoxicity** | kg 1,4-DCB | 0.000941 | 0.000562 | 5.64E-06 | 6.21E-06 | 3.61E-06 | 2.06E-05 | 5.41E-06 | 0.000165 | 0.000172 |
| **Marine ecotoxicity** | kg 1,4-DCB | 0.001255 | 0.000744 | 7.39E-06 | 8.28E-06 | 6.29E-06 | 2.73E-05 | 7.72E-06 | 0.000221 | 0.000233 |
| **Human carcinogenic toxicity** | kg 1,4-DCB | 0.001555 | 0.000359 | 3.78E-06 | 7.11E-06 | 3.54E-06 | 9.48E-06 | 1.14E-05 | 0.000272 | 0.000889 |
| **Human non-carcinogenic toxicity** | kg 1,4-DCB | 0.023014 | 0.012482 | 0.000137 | 0.000195 | 0.000119 | 0.000417 | 9.00E-05 | 0.005207 | 0.004367 |
| **Land use** | m^2^a crop eq | 0.000582 | 0.000298 | 5.71E-05 | 8.33E-05 | 8.42E-06 | 5.74E-06 | 1.79E-06 | 6.69E-05 | 6.11E-05 |
| **Mineral resource scarcity** | kg Cu eq | 0.000288 | 0.000273 | 3.72E-07 | 5.44E-07 | 5.76E-07 | 1.01E-06 | 1.27E-06 | 3.37E-06 | 7.20E-06 |
| **Fossil resource scarcity** | kg oil eq | 0.008677 | 0.005599 | 3.59E-05 | 6.13E-05 | 5.32E-05 | 6.90E-05 | 0.000149 | 0.001998 | 0.000712 |
| **Water consumption** | m^3^ | 0.000232 | 0.000147 | 1.52E-06 | 2.91E-06 | 2.84E-07 | 5.21E-07 | 3.36E-07 | 5.18E-05 | 2.81E-05 |

Environmental impacts (midpoint categories) measured using life cycle assessment modelled on one glove (with shipping, and waste via hazardous incineration). 1,4-DCB =dichlorobenzene, CFC11= Trichlorofluoromethane, CO_2_e= carbon dioxide equivalents, Cu= copper, eq= equivalents, kBq Co-60 eq = kilobecquerel Cobalt-60, m^2^a = square meter years, N= nitrogen, NO_x_= nitrous oxides, P=phosphate, PM2.5 = particulate matter <2.5 micrometers, SO_2_= sulphur dioxide

**Supplementary Table 9: Environmental impact of one disposable gown**

| **Impact category** | **Unit** | **Total** | Textile, non-woven polypropylene {GLO}\| market for textile, non woven polypropylene \| Cut-off, U | Synthetic rubber {GLO}\| market for \| Cut-off, U | Packaging film, low density polyethylene {GLO}\| market for \| Cut-off, U | Kraft paper, bleached {GLO}\| market for \| Cut-off, U | Corrugated board box {RoW}\| market for corrugated board box \| Cut-off, U | Transport, freight, sea, container ship {GLO}\| market for transport, freight, sea, container ship \| Cut-off, S | Transport, freight, light commercial vehicle {GLO}\| market group for transport, freight, light commercial vehicle \| Cut-off, U | Transport, freight, lorry, unspecified {GLO}\| market group for transport, freight, lorry, unspecified \| Cut-off, U | Electricity, medium voltage {CN}\| market group for \| Cut-off, U | Electricity, medium voltage {EG}\| market for electricity, medium voltage \| Cut-off, U | Electricity, medium voltage {DE}\| market for \| Cut-off, U | PPE clinical waste |
| --- | --- | --- | --- | --- | --- | --- | --- | --- | --- | --- | --- | --- | --- | --- |
| **Global warming** | kg CO_2_e | 0.905431 | 0.404429 | 0.021734 | 0.014277 | 0.00956 | 0.012208 | 0.020126 | 0.010184 | 0.007387 | 0.026729 | 0.006958 | 0.005253 | 0.366184 |
| **Stratospheric ozone depletion** | kg CFC11 eq | 3.18E-07 | 1.16E-07 | 1.39E-08 | 3.62E-09 | 5.42E-09 | 1.22E-08 | 1.41E-08 | 6.15E-09 | 3.46E-09 | 5.70E-09 | 3.44E-09 | 3.71E-09 | 1.30E-07 |
| **Ionizing radiation** | kBq Co-60 eq | 0.034539 | 0.023092 | 0.001713 | 0.000899 | 0.000851 | 0.000364 | 0.000209 | 0.000316 | 0.000144 | 0.00041 | 2.61E-05 | 0.000954 | 0.005564 |
| **Ozone formation, Human health** | kg NO_x_ eq | 0.00204 | 0.000959 | 5.68E-05 | 3.45E-05 | 3.26E-05 | 3.82E-05 | 0.000419 | 4.57E-05 | 4.01E-05 | 7.40E-05 | 1.13E-05 | 5.42E-06 | 0.000323 |
| **Fine particulate matter formation** | kg PM2.5 eq | 0.001098 | 0.000538 | 3.87E-05 | 1.92E-05 | 2.63E-05 | 2.01E-05 | 0.000134 | 1.62E-05 | 1.08E-05 | 4.08E-05 | 5.35E-06 | 2.78E-06 | 2.45E-04 |
| **Ozone formation, Terrestrial ecosystems** | kg NOx eq | 0.002107 | 0.001005 | 6.16E-05 | 3.73E-05 | 3.32E-05 | 3.89E-05 | 0.000422 | 4.71E-05 | 4.09E-05 | 7.42E-05 | 1.15E-05 | 5.48E-06 | 0.000329 |
| **Terrestrial acidification** | kg SO_2_ eq | 0.002644 | 0.001212 | 8.41E-05 | 4.23E-05 | 3.45E-05 | 4.31E-05 | 0.000415 | 3.53E-05 | 2.48E-05 | 9.11E-05 | 1.65E-05 | 9.02E-06 | 0.000634 |
| **Freshwater eutrophication** | kg P eq | 0.000272 | 0.000109 | 6.85E-06 | 3.99E-06 | 3.79E-06 | 4.85E-06 | 7.91E-07 | 1.89E-06 | 6.79E-07 | 4.88E-06 | 1.18E-07 | 7.41E-06 | 1.28E-04 |
| **Marine eutrophication** | kg N eq | 2.25E-05 | 9.84E-06 | 4.96E-07 | 4.89E-07 | 5.29E-07 | 2.76E-06 | 7.60E-08 | 1.14E-07 | 5.42E-08 | 3.16E-07 | 1.06E-08 | 4.74E-07 | 7.39E-06 |
| **Terrestrial ecotoxicity** | kg 1,4-DCB | 1.849938 | 0.942835 | 0.080409 | 0.029279 | 0.031426 | 0.051025 | 0.053655 | 0.067043 | 0.141189 | 0.017658 | 0.010597 | 0.003274 | 0.418905 |
| **Freshwater ecotoxicity** | kg 1,4-DCB | 0.028206 | 0.015775 | 0.001346 | 0.000455 | 0.000312 | 0.000541 | 0.000216 | 0.000989 | 0.000173 | 0.000411 | 8.79E-05 | 0.000257 | 0.007606 |
| **Marine ecotoxicity** | kg 1,4-DCB | 0.037237 | 0.020456 | 0.001781 | 0.000599 | 0.000417 | 0.000709 | 0.000308 | 0.001309 | 0.000302 | 0.000543 | 0.000115 | 0.000343 | 0.010304 |
| **Human carcinogenic toxicity** | kg 1,4-DCB | 0.056156 | 0.012537 | 0.000859 | 0.000443 | 0.000415 | 0.000362 | 0.000454 | 0.000455 | 0.00017 | 0.000751 | 3.72E-05 | 0.000378 | 0.03928 |
| **Human non-carcinogenic toxicity** | kg 1,4-DCB | 0.583373 | 0.278272 | 0.02988 | 0.00985 | 0.007697 | 0.013184 | 0.003587 | 0.019995 | 0.00569 | 0.012643 | 0.000802 | 0.008112 | 0.1929 |
| **Land use** | m^2^a crop eq | 0.024286 | 0.004788 | 0.000713 | 0.000467 | 0.008952 | 0.005479 | 7.16E-05 | 0.000275 | 0.000404 | 0.00032 | 9.69E-06 | 9.96E-05 | 0.002698 |
| **Mineral resource scarcity** | kg Cu eq | 0.002298 | 0.001086 | 6.54E-04 | 2.96E-05 | 2.65E-05 | 3.57E-05 | 5.08E-05 | 4.86E-05 | 2.76E-05 | 1.11E-05 | 3.34E-06 | 4.86E-06 | 0.000318 |
| **Fossil resource scarcity** | kg oil eq | 0.335342 | 0.254846 | 0.013402 | 0.008436 | 0.002754 | 0.00344 | 0.005939 | 0.003314 | 0.002551 | 0.00519 | 0.002625 | 0.001263 | 0.03145 |
| **Water consumption** | m^3^ | 0.006301 | 0.003863 | 0.000351 | 0.000243 | 0.000296 | 0.000146 | 1.34E-05 | 2.50E-05 | 1.36E-05 | 6.57E-05 | 1.41E-05 | 2.94E-05 | 0.00124 |

Environmental impacts (midpoint categories) measured using life cycle assessment modelled on one gown (with shipping, and waste via hazardous incineration). 1,4-DCB =dichlorobenzene, CFC11= Trichlorofluoromethane, CO_2_e= carbon dioxide equivalents, Cu= copper, eq= equivalents, kBq Co-60 eq = kilobecquerel Cobalt-60, m^2^a = square meter years, N= nitrogen, NO_x_= nitrous oxides, P=phosphate, PM2.5 = particulate matter <2.5 micrometers, SO_2_= sulphur dioxide

**Supplementary Table 10: Environmental impact of one surgical mask (Type II)**

| **Impact category** | **Unit** | **Total** | Textile, non-woven polypropylene {GLO}\| market for textile, non woven polypropylene \| Cut-off, U | Synthetic rubber {GLO}\| market for \| Cut-off, U | Sheet rolling, aluminium {GLO}\| market for \| Cut-off, U | Corrugated board box {RoW}\| market for corrugated board box \| Cut-off, U | Folding boxboard/chipboard {GLO}\| market for \| Cut-off, U | Transport, freight, lorry, unspecified {GLO}\| market group for transport, freight, lorry, unspecified \| Cut-off, U | Transport, freight, light commercial vehicle {GLO}\| market group for transport, freight, light commercial vehicle \| Cut-off, U | Transport, freight, sea, container ship {GLO}\| market for transport, freight, sea, container ship \| Cut-off, U | Electricity, medium voltage {CN}\| market group for \| Cut-off, U | Electricity, medium voltage {GB}\| market for \| Cut-off, U | Electricity, medium voltage {MX}\| market for \| Cut-off, U | PPE clinical waste |
| --- | --- | --- | --- | --- | --- | --- | --- | --- | --- | --- | --- | --- | --- | --- |
| **Global warming** | kg CO_2_e | 0.013491 | 0.004502 | 0.000788 | 0.000151 | 0.000281 | 0.001226 | 0.000141 | 0.000194 | 0.000492 | 0.000493 | 3.20E-05 | 2.51E-05 | 0.005167 |
| **Stratospheric ozone depletion** | kg CFC11 eq | 5.25E-09 | 1.29E-09 | 5.02E-10 | 5.26E-11 | 2.80E-10 | 6.10E-10 | 6.58E-11 | 1.17E-10 | 3.45E-10 | 1.05E-10 | 1.64E-11 | 3.32E-11 | 1.84E-09 |
| **Ionizing radiation** | kBq Co-60 eq | 0.000545 | 0.000257 | 6.21E-05 | 1.12E-05 | 8.39E-06 | 8.61E-05 | 2.75E-06 | 6.01E-06 | 5.11E-06 | 7.56E-06 | 1.84E-05 | 1.51E-06 | 7.85E-05 |
| **Ozone formation, Human health** | kg NO_x_ eq | 3.52E-05 | 1.07E-05 | 2.06E-06 | 3.42E-07 | 8.80E-07 | 3.33E-06 | 7.64E-07 | 8.70E-07 | 1.02E-05 | 1.37E-06 | 5.59E-08 | 4.29E-08 | 4.55E-06 |
| **Fine particulate matter formation** | kg PM2.5 eq | 1.86E-05 | 5.99E-06 | 1.40E-06 | 2.88E-07 | 4.62E-07 | 2.35E-06 | 2.05E-07 | 3.09E-07 | 3.27E-06 | 7.53E-07 | 2.59E-08 | 5.78E-08 | 3.46E-06 |
| **Ozone formation, Terrestrial ecosystems** | kg NOx eq | 3.61E-05 | 1.12E-05 | 2.23E-06 | 3.59E-07 | 8.98E-07 | 3.37E-06 | 7.79E-07 | 8.97E-07 | 1.03E-05 | 1.37E-06 | 5.65E-08 | 4.36E-08 | 4.64E-06 |
| **Terrestrial acidification** | kg SO_2_ eq | 4.50E-05 | 1.35E-05 | 3.05E-06 | 5.05E-07 | 9.94E-07 | 4.91E-06 | 4.73E-07 | 6.73E-07 | 1.01E-05 | 1.68E-06 | 7.54E-08 | 8.24E-08 | 8.95E-06 |
| **Freshwater eutrophication** | kg P eq | 4.04E-06 | 1.21E-06 | 2.48E-07 | 5.89E-08 | 1.12E-07 | 4.29E-07 | 1.29E-08 | 3.59E-08 | 1.93E-08 | 9.00E-08 | 6.73E-09 | 1.13E-08 | 1.80E-06 |
| **Marine eutrophication** | kg N eq | 3.95E-07 | 1.10E-07 | 1.80E-08 | 4.97E-09 | 6.36E-08 | 8.24E-08 | 1.03E-09 | 2.16E-09 | 1.86E-09 | 5.82E-09 | 7.58E-10 | 7.39E-10 | 1.04E-07 |
| **Terrestrial ecotoxicity** | kg 1,4-DCB | 0.029517 | 0.010496 | 0.002916 | 0.000173 | 0.001176 | 0.00318 | 0.002688 | 0.001276 | 0.00131 | 0.000326 | 3.48E-05 | 2.84E-05 | 0.00591 |
| **Freshwater ecotoxicity** | kg 1,4-DCB | 0.00042 | 0.000176 | 4.88E-05 | 6.83E-06 | 1.25E-05 | 3.26E-05 | 3.30E-06 | 1.88E-05 | 5.26E-06 | 7.57E-06 | 8.48E-07 | 5.87E-07 | 0.000107 |
| **Marine ecotoxicity** | kg 1,4-DCB | 0.000556 | 0.000228 | 6.46E-05 | 8.81E-06 | 1.63E-05 | 4.35E-05 | 5.75E-06 | 2.49E-05 | 7.52E-06 | 1.00E-05 | 1.15E-06 | 7.82E-07 | 0.000145 |
| **Human carcinogenic toxicity** | kg 1,4-DCB | 0.00082 | 0.00014 | 3.11E-05 | 1.12E-05 | 8.35E-06 | 3.73E-05 | 3.24E-06 | 8.67E-06 | 1.11E-05 | 1.38E-05 | 7.55E-07 | 6.94E-07 | 0.000554 |
| **Human non-carcinogenic toxicity** | kg 1,4-DCB | 0.009203 | 0.003098 | 0.001084 | 0.000129 | 0.000304 | 0.001023 | 0.000108 | 0.000381 | 8.76E-05 | 0.000233 | 1.91E-05 | 1.45E-05 | 0.002722 |
| **Land use** | m^2^a crop eq | 0.000707 | 5.33E-05 | 2.59E-05 | 1.92E-06 | 0.000126 | 0.000437 | 7.70E-06 | 5.24E-06 | 1.75E-06 | 5.91E-06 | 3.55E-06 | 4.71E-08 | 3.81E-05 |
| **Mineral resource scarcity** | kg Cu eq | 4.75E-05 | 1.21E-05 | 2.37E-05 | 5.70E-07 | 8.23E-07 | 2.86E-06 | 5.26E-07 | 9.25E-07 | 1.24E-06 | 2.05E-07 | 4.04E-08 | 1.31E-08 | 4.49E-06 |
| **Fossil resource scarcity** | kg oil eq | 0.004577 | 0.002837 | 0.000486 | 3.86E-05 | 7.93E-05 | 0.000322 | 4.86E-05 | 6.31E-05 | 0.000145 | 9.57E-05 | 1.08E-05 | 7.69E-06 | 0.000444 |
| **Water consumption** | m^3^ | 9.55E-05 | 4.30E-05 | 1.27E-05 | 1.17E-06 | 3.36E-06 | 1.53E-05 | 2.59E-07 | 4.76E-07 | 3.28E-07 | 1.21E-06 | 1.40E-07 | 4.80E-08 | 1.75E-05 |

Environmental impacts (midpoint categories) measured using life cycle assessment modelled on one Type II surgical mask (with shipping, and waste via hazardous incineration). 1,4-DCB =dichlorobenzene, CFC11= Trichlorofluoromethane, CO_2_e= carbon dioxide equivalents, Cu= copper, eq= equivalents, kBq Co-60 eq = kilobecquerel Cobalt-60, m^2^a = square meter years, N= nitrogen, NO_x_= nitrous oxides, P=phosphate, PM2.5 = particulate matter <2.5 micrometers, SO_2_= sulphur dioxide

**Supplementary Table 11: Environmental impact of one surgical mask (Type IIR)**

| **Impact category** | **Unit** | **Total** | Textile, non-woven polypropylene {GLO}\| market for textile, non woven polypropylene \| Cut-off, U | Sheet rolling, aluminium {GLO}\| market for \| Cut-off, U | Synthetic rubber {GLO}\| market for \| Cut-off, U | Corrugated board box {RoW}\| market for corrugated board box \| Cut-off, U | Folding boxboard/chipboard {GLO}\| market for \| Cut-off, U | T Transport, freight, sea, container ship {GLO}\| market for transport, freight, sea, container ship \| Cut-off, U | Transport, freight, light commercial vehicle {GLO}\| market group for transport, freight, light commercial vehicle \| Cut-off, U | Transport, freight, lorry, unspecified {GLO}\| market group for transport, freight, lorry, unspecified \| Cut-off, U | Electricity, medium voltage {CN}\| market group for \| Cut-off, U | Electricity, medium voltage {MX}\| market for \| Cut-off, U | Electricity, medium voltage {GB}\| market for \| Cut-off, U | PPE clinical waste |
| --- | --- | --- | --- | --- | --- | --- | --- | --- | --- | --- | --- | --- | --- | --- |
| **Global warming** | kg CO_2_e | 0.020192 | 0.007354 | 0.000131 | 0.001267 | 0.000302 | 0.00124 | 0.000656 | 0.000259 | 0.000188 | 0.00076 | 3.87E-05 | 7.40E-05 | 0.007968 |
| **Stratospheric ozone depletion** | kg CFC11 eq | 7.63E-09 | 2.10E-09 | 4.57E-11 | 8.07E-10 | 3.00E-10 | 6.17E-10 | 4.60E-10 | 1.56E-10 | 8.78E-11 | 1.62E-10 | 5.13E-11 | 3.80E-11 | 2.83E-09 |
| **Ionizing radiation** | kBq Co-60 eq | 0.000807 | 0.00042 | 9.76E-06 | 9.98E-05 | 8.99E-06 | 8.71E-05 | 6.81E-06 | 8.02E-06 | 3.66E-06 | 1.17E-05 | 2.33E-06 | 4.26E-05 | 1.21E-04 |
| **Ozone formation, Human health** | kg NO_x_ eq | 5.04E-05 | 1.74E-05 | 2.98E-07 | 3.31E-06 | 9.42E-07 | 3.37E-06 | 1.36E-05 | 1.16E-06 | 1.02E-06 | 2.11E-06 | 6.61E-08 | 1.29E-07 | 7.02E-06 |
| **Fine particulate matter formation** | kg PM2.5 eq | 2.68E-05 | 9.78E-06 | 2.50E-07 | 2.26E-06 | 4.95E-07 | 2.37E-06 | 4.35E-06 | 4.13E-07 | 2.73E-07 | 1.16E-06 | 8.92E-08 | 6.00E-08 | 5.33E-06 |
| **Ozone formation, Terrestrial ecosystems** | kg NOx eq | 5.19E-05 | 1.83E-05 | 3.12E-07 | 3.59E-06 | 9.62E-07 | 3.41E-06 | 1.37E-05 | 1.20E-06 | 1.04E-06 | 2.11E-06 | 6.73E-08 | 1.31E-07 | 7.15E-06 |
| **Terrestrial acidification** | kg SO_2_ eq | 6.50E-05 | 2.20E-05 | 4.39E-07 | 4.90E-06 | 1.06E-06 | 4.97E-06 | 1.35E-05 | 8.97E-07 | 6.30E-07 | 2.59E-06 | 1.27E-07 | 1.74E-07 | 1.38E-05 |
| **Freshwater eutrophication** | kg P eq | 6.01E-06 | 1.98E-06 | 5.12E-08 | 3.99E-07 | 1.20E-07 | 4.34E-07 | 2.58E-08 | 4.79E-08 | 1.72E-08 | 1.39E-07 | 1.75E-08 | 1.56E-08 | 2.78E-06 |
| **Marine eutrophication** | kg N eq | 5.38E-07 | 1.79E-07 | 4.32E-09 | 2.89E-08 | 6.82E-08 | 8.34E-08 | 2.48E-09 | 2.89E-09 | 1.38E-09 | 8.98E-09 | 1.14E-09 | 1.75E-09 | 1.61E-07 |
| **Terrestrial ecotoxicity** | kg 1,4-DCB | 0.043126 | 0.017144 | 0.000151 | 0.004687 | 0.00126 | 0.003218 | 0.001748 | 0.001702 | 0.003585 | 0.000502 | 4.38E-05 | 8.06E-05 | 0.009116 |
| **Freshwater ecotoxicity** | kg 1,4-DCB | 0.000633 | 0.000287 | 5.94E-06 | 7.85E-05 | 1.34E-05 | 3.30E-05 | 7.02E-06 | 2.51E-05 | 4.40E-06 | 1.17E-05 | 9.05E-07 | 1.96E-06 | 0.000166 |
| **Marine ecotoxicity** | kg 1,4-DCB | 0.000837 | 0.000372 | 7.66E-06 | 0.000104 | 1.75E-05 | 4.40E-05 | 1.00E-05 | 3.32E-05 | 7.66E-06 | 1.54E-05 | 1.21E-06 | 2.67E-06 | 0.000224 |
| **Human carcinogenic toxicity** | kg 1,4-DCB | 0.001243 | 0.000228 | 9.70E-06 | 5.01E-05 | 8.94E-06 | 3.78E-05 | 1.48E-05 | 1.16E-05 | 4.32E-06 | 2.14E-05 | 1.07E-06 | 1.75E-06 | 0.000855 |
| **Human non-carcinogenic toxicity** | kg 1,4-DCB | 0.013631 | 0.00506 | 0.000113 | 0.001742 | 0.000326 | 0.001035 | 0.000117 | 0.000508 | 0.000144 | 0.00036 | 2.24E-05 | 4.41E-05 | 0.004198 |
| **Land use** | m^2^a crop eq | 0.000792 | 8.71E-05 | 1.67E-06 | 4.16E-05 | 0.000135 | 0.000442 | 2.33E-06 | 6.99E-06 | 1.03E-05 | 9.11E-06 | 7.26E-08 | 8.20E-06 | 5.87E-05 |
| **Mineral resource scarcity** | kg Cu eq | 7.30E-05 | 1.97E-05 | 4.96E-07 | 3.81E-05 | 8.82E-07 | 2.89E-06 | 1.65E-06 | 1.23E-06 | 7.02E-07 | 3.17E-07 | 2.02E-08 | 9.34E-08 | 6.92E-06 |
| **Fossil resource scarcity** | kg oil eq | 0.007056 | 0.004634 | 3.36E-05 | 0.000781 | 8.50E-05 | 0.000326 | 0.000193 | 8.41E-05 | 6.48E-05 | 0.000148 | 1.19E-05 | 2.51E-05 | 0.000684 |
| **Water consumption** | m^3^ | 0.000141 | 7.02E-05 | 1.02E-06 | 2.05E-05 | 3.60E-06 | 1.55E-05 | 4.37E-07 | 6.35E-07 | 3.46E-07 | 1.87E-06 | 7.40E-08 | 3.24E-07 | 2.70E-05 |

Environmental impacts (midpoint categories) measured using life cycle assessment modelled on one Type IIR surgical mask (with shipping, and waste via hazardous incineration). 1,4-DCB =dichlorobenzene, CFC11= Trichlorofluoromethane, CO_2_e= carbon dioxide equivalents, Cu= copper, eq= equivalents, kBq Co-60 eq = kilobecquerel Cobalt-60, m^2^a = square meter years, N= nitrogen, NO_x_= nitrous oxides, P=phosphate, PM2.5 = particulate matter <2.5 micrometers, SO_2_= sulphur dioxide

**Supplementary Table 12: Environmental impact of scenario models (midpoint categories)**

| **Impact category** | **Unit** | Shipping  (Base scenario) | Air freight | UK manufacture | Reduce | Reuse | Recycle | Combination |
| --- | --- | --- | --- | --- | --- | --- | --- | --- |
| **Global warming** | kg CO_2_e | 1.06E+08 | 1.59E+08 | 9.40E+07 | 5.82E+07 | 9.54E+07 | 6.92E+07 | 2.66E+07 |
| **Stratospheric ozone depletion** | kg CFC11 eq | 40.538965 | 52.84834 | 37.8953 | 19.1113 | 37.169319 | 27.297802 | 8.5314768 |
| **Ionizing radiation** | kBq Co-60 eq | 3882726.8 | 4393781 | 8070889 | 2247673 | 3586704.4 | 3316442 | 2191322.8 |
| **Ozone formation, Human health** | kg NO_x_ eq | 244352.67 | 467441 | 172731 | 140816.6 | 216256.7 | 211507.29 | 64913.055 |
| **Fine particulate matter formation** | kg PM2.5 eq | 157695.7 | 195349.3 | 106342.3 | 74078.88 | 143214.97 | 132765.47 | 36093.199 |
| **Ozone formation, Terrestrial ecosystems** | kg NOx eq | 254583.89 | 479487.2 | 182411.1 | 146581.1 | 225403.83 | 221150.88 | 69118.6 |
| **Terrestrial acidification** | kg SO_2_ eq | 341021.9 | 452932.5 | 247915.7 | 179785.7 | 305172.63 | 276487.71 | 80324.334 |
| **Freshwater eutrophication** | kg P eq | 33786.378 | 34486.74 | 28534.56 | 16823.66 | 30591.98 | 20784.428 | 7122.7384 |
| **Marine eutrophication** | kg N eq | 3017.4222 | 3071.634 | 2739.373 | 1838.291 | 2720.1492 | 2265.1849 | 1169.83 |
| **Terrestrial ecotoxicity** | kg 1,4-DCB | 2.11E+08 | 2.97E+08 | 2.01E+08 | 1.10E+08 | 1.90E+08 | 1.68E+08 | 65225857 |
| **Freshwater ecotoxicity** | kg 1,4-DCB | 3361093.7 | 3507965 | 3174324 | 1630832 | 3036956.3 | 2587047.1 | 912910.91 |
| **Marine ecotoxicity** | kg 1,4-DCB | 4475710.3 | 4726242 | 4228065 | 2167458 | 4045755.3 | 3427117.7 | 1204522.6 |
| **Human carcinogenic toxicity** | kg 1,4-DCB | 6345112.4 | 6470094 | 5867629 | 3484960 | 5646296.9 | 2347669.2 | 829587.02 |
| **Human non-carcinogenic toxicity** | kg 1,4-DCB | 78904366 | 89983829 | 71674682 | 36576646 | 7.20E+07 | 59273197 | 19548776 |
| **Land use** | m^2^a crop eq | 2634024.8 | 2728783 | 3295467 | 1563170 | 2640316.1 | 2359449.1 | 1518460.9 |
| **Mineral resource scarcity** | kg Cu eq | 667193.13 | 679485.6 | 662431.3 | 138218 | 638533.59 | 634807.18 | 91173.241 |
| **Fossil resource scarcity** | kg oil eq | 36593143 | 53682001 | 34013493 | 20633254 | 32450424 | 33392554 | 14406081 |
| **Water consumption** | m^3^ | 973330.73 | 992483.5 | 900477.8 | 546163.7 | 863685.01 | 847130.81 | 373862 |

Environmental impact (midpoint categories) of PPE distributed to health and social care services in England by NHS Supply Chain between 25^th^ February and 23^rd^ August 2020,, measured using life cycle assessment, modelling base scenario (shipping, single-use PPE, clinical waste), air freight, UK manufacture, reduce (zero glove use), reuse (reusable gown, reuse of face shield, all other items single-use), recycling, and combination of measures. 1,4-DCB =dichlorobenzene, CFC11= Trichlorofluoromethane, CO_2_e= carbon dioxide equivalents, Cu= copper, eq= equivalents, kBq Co-60 eq = kilobecquerel Cobalt-60, m^2^a = square meter years, N= nitrogen, NO_x_= nitrous oxides, P=phosphate, PM2.5 = particulate matter <2.5 micrometers, SO_2_= sulphur dioxide

**Supplementary Table 13: Environmental impact of scenario models (endpoint categories)**

| **Damage category** | **Unit** | Shipping  (Base Scenario) | Air freight | UK manufacture | Reduce | Reuse | Recycle | Combination |
| --- | --- | --- | --- | --- | --- | --- | --- | --- |
| **Human health** | DALY | 239 | 314 | 192 | 122 | 215 | 171 | 55 |
| **Ecosystems** | species.yr | 0.466 | 0.668 | 0.403 | 0.255 | 0.419 | 0.330 | 0.125 |
| **Resources** | US $ | 12,697,261 | 20,366,321 | 12,253,727 | 7,277,962 | 11,188,442 | 12,018,323 | 5,320,902 |

Environmental impact (endpoint categories) of PPE distributed to health and social care services in England by NHS Supply Chain between 25^th^ February and 23^rd^ August 2020, measured using life cycle assessment, modelling base scenario (shipping, single-use PPE, clinical waste), air freight, UK manufacture, reduce (zero glove use), reuse (reusable gown, reuse of face shield, all other items single-use), recycling, and combination of measures (UK manufacture, reduce, reuse and recycle). DALYs= disability adjusted life years, species.year=loss of local species per year, US$=extra costs involved for future mineral and fossil resource extraction
